# Context-dependent associations between recent antibiotic treatment, gut microbiome composition, blood metabolites, and metabolic health

**DOI:** 10.64898/2026.11.19.26368536

**Authors:** Crystal Perez, Sarah K. Alver, Lance Pflieger, Nick Quinn-Bohmann, Noa Rappaport, Andrew T. Magis, Robert D. Burk, Qibin Qi, Robert Kaplan, Sean M. Gibbons

**Affiliations:** Institute for Systems Biology, Seattle, WA 98109, USA; Molecular Engineering Graduate Program, University of Washington, Seattle, WA 98195, USA; Medical Scientist Training Program, University of Washington, Seattle, WA 98195, USA; Department of Epidemiology, Fred Hutchinson Cancer Center, Seattle, WA 98109, USA; Phenome Health, Seattle, WA 98121, USA; Buck Institute for Research on Aging, Novato 94945, CA, USA; Department of Epidemiology & Population Health, Albert Einstein College of Medicine, Bronx, NY 10461, USA; Department of Biomedical Engineering, Tandon School of Engineering, New York University, Brooklyn, NY 11201; Department of Bioengineering, University of Washington, Seattle, WA 98195, USA; Department of Genome Sciences, University of Washington, Seattle, WA 98195, USA; Department of Environmental and Occupational Health Sciences, University of Washington, Seattle, WA 98195, USA

## Abstract

Antibiotics are among the most widely prescribed drugs, and they reliably disrupt the commensal gut microbiota. Large epidemiological studies link antibiotic exposure to type 2 diabetes (T2D) risk, yet smaller randomized intervention trials have failed to find this connection. Whether this discordance reflects a true lack of signal or subpopulation-dependent responses to antibiotics remains unclear. Here, across three independent cohorts (N=4,122), we show that recent antibiotic use may reshape host physiology in specific dietary, microbial, and metabolic contexts. Recent antibiotic use was associated with reduced gut microbiome richness, greater *Bacteroides* and pathobiont dominance, lower predicted butyrate production, and altered levels of 34 blood metabolites, including several microbe–host co-metabolites, such as hippurate and isoursodeoxycholic acid, while the blood proteome was unchanged. Diet and microbiome composition influenced the magnitude of these effects: preference for liquor was associated with steeper diversity loss, and the presence of specific bacterial genera either dampened or amplified antibiotic-associated blood metabolite differences. Clinical chemistries did not differ, on average, across antibiotic treament groups, affirming the intervention trials. However, recent antibiotic users with T2D showed elevated methylmalonic acid, homocysteine, and alkaline phosphatase, suggesting B-vitamin deficiency. Furthermore, antibiotic users with poorer metabolic health (e.g., blood metabolomics-inferredBMI > measured BMI) showed lower circulating 25-hydroxyvitamin D. Overall, metabolic disruption associated with recent antibiotic use appears to be related to vitamin deficiencies and is concentrated in metabolically vulnerable subgroups, arguing for further research into context-aware prescribing and targeted co-therapies, like higher fiber diets, probiotics, or supplements.

## Introduction

Fecal metagenomic and 16S amplicon sequencing analyses have revealed that antibiotic treatment can promote microbiome phenotypes mirroring those of individuals with cardiometabolic disease or T2D ^1,2^. Similarly, antibiotic exposure has been linked to increased risk of developing Type 2 Diabetes (T2D) in epidemiological studies that leverage either electronic health records ^3–10^ or environmental monitoring ^11,12^. These results are, perhaps, unsurprising, given that the gut microbiota is known to contribute to host energy balance ^13,14^ and has been widely associated with a range of obesity-related phenotypes ^13,15–17^. However, epidemiological studies are limited by unmeasured confounders and lack the causal certainty of a randomized intervention trial.

Most epidemiological studies linking antibiotic use with T2D risk show a positive dose-response association, yet adjustment for confounding variables appears to ablate this association. For example, a nested case-control study from Alberta’s Tomorrow Project found no association between antibiotic use and T2D after controlling for self-reported clinical and lifestyle measures (e.g., socioeconomic status, body mass index (BMI), leisure time physical activity and overall diet quality) that are not always captured in other observational cohorts ^5^. This finding is consistent with smaller, randomized, placebo-controlled intervention studies that have not found lasting impacts on host metabolism following antibiotic treatment ^18,19^. In aggregate, the existing literature suggests that there is a need to collect more extensive molecular phenotype data in addition to other lifestyle and dietary metadata in antibiotic users, as the putative metabolic risk of antibiotic treatment may vary with population- or individual-level variables ^5,20–23^.

Antibiotic-induced perturbations of the gut microbiota include transient declines in total bacterial biomass, changes in community composition, and incomplete ecological recovery over time^24–26^. Indeed, a recent epidemiological study linked gut microbiome samples from three Swedish cohorts to the Swedish National Prescribed Drug Register and detected persistent ecological effects of certain antibiotic classes, such as fluoroquinolones, up to eight years post-treatment.^2^ The ecological resistance and resilience of the gut microbiota to antibiotic treatment can vary across individuals and appears to be related to baseline microbiome composition, dietary intake, and host behavior ^27–29^. Prior work from our group and from others has found an intimate connection between human gut microbiome composition and the abundances of circulating metabolites in blood ^30–32^, which aligns with antibiotic treatment studies showing how antibiotics can affect the blood metabolome. For example, a recent study demonstrated that antibiotic use strongly impacts the fecal metabolome and modestly influences plasma metabolites ^33,34^. Likewise, lower serum levels of five metabolites—all of which can involve bacterial metabolism ^35–37^—were reported in 20 hospitalized patients who received antibiotics, compared to five hospitalized controls ^38^.

Many interventional studies have investigated the impact of various antibiotic classes on the commensal gut microbiota in humans and rodents ^18,19,39–41^. However, these studies often have small sample sizes, limiting their statistical power and generalizability. In contrast, for many large, cross-sectional cohort studies that collect stool and blood samples for multi-omic analyses, recent antibiotic use is treated as an exclusion criterion, rather than as an exposure of interest. Baldanzi et al. clearly demonstrated the value of linking prescription drug information to microbiome data in order to learn about the long-term consequences of antibiotics on the gut microbiome, but they lacked paired host molecular phenotype data ^2^. The broader consequences of recent antibiotic use on host physiology, as assessed through dense multi-omic measurements and clinical chemistries, across thousands of individuals with different dietary, lifestyle, and gut microbiota contexts, remain poorly understood.

Here, we used data from 4,122 individuals across three large, deeply phenotyped populations to characterize the context-dependent impacts of recent antibiotic use on the host molecular phenotype by integrating gut microbiome, blood metabolome, blood proteome, clinical chemistry, and questionnaire data on diet, demographics, and behavior.

## Results

### Study design and cohort composition

We used data from two United States-based cohorts and one European cohort: Arivale (n = 2,666) ^42^, Hispanic Community Health Study/Study of Latinos (SOL; n = 866) ^43^, and MetaCardis (MC; n = 590) ^1,44^, respectively. The Arivale cohort consists primarily of non-Hispanic, white adults who were formerly subscribers of the Arivale Scientific Wellness program.^45,46^ Individuals in Arivale for whom antibiotic usage information was available were eligible for analysis (n = 3,155), resulting in a cohort of 2,666 after excluding participants with missing covariate data (Fig. S1, methods). We validated our Arivale findings using data from HCHS/SOL and MetaCardis. The HCHS/SOL cohort consists of 16,415 Hispanic adults (US-born and non-US-born) residing in the US; analyses were restricted to those with antibiotic usage and multi-omic data (N=866) whose clinical data were collected within 60 days of the stool sample. Finally, the MC cohort includes European individuals across the spectrum of cardiometabolic disease. For this study, we analyzed MC cohort data of individuals without heart disease and available fecal metagenomes, blood metabolomes, antibiotic use data (N=590).

Antibiotic use metadata were operationalized differently across the three cohorts; Arivale and HCHS/SOL use time since last antibiotic use with cutoffs at 12 or 6 months (Fig. 1A, Table S1), while MC counted the number of antibiotic courses in the five years prior to sampling. Importantly, antibiotic recency groups represent broad windows, so time since last antibiotic use is unresolved within each group (e.g., within the 3-month group, an individual sampled 10 days after antibiotic use is grouped with one sampled 90 days after). Moreover, our exposure includes the association for both the antibiotic and the treated condition, which we cannot distinguish in this study. Antibiotic use within the past three months or within the past twelve months was reported by 428 (16.1%) and 593 (22.2%) Arivale participants, respectively, with the remaining 1,645 (61.7%) reporting no antibiotic use in the last year. In HCHS/SOL, 136 (15.0%) reported use within three months and 72 (8.3%) within the previous 6 months. Because analyses were restricted to participants whose samples were collected within 30 days (Arivale) or 60 days (SOL) of antibiotic recency ascertainment, per-assay sample sizes are smaller than the full cohorts and vary by data type (Fig. 1A).

**Figure 1.**
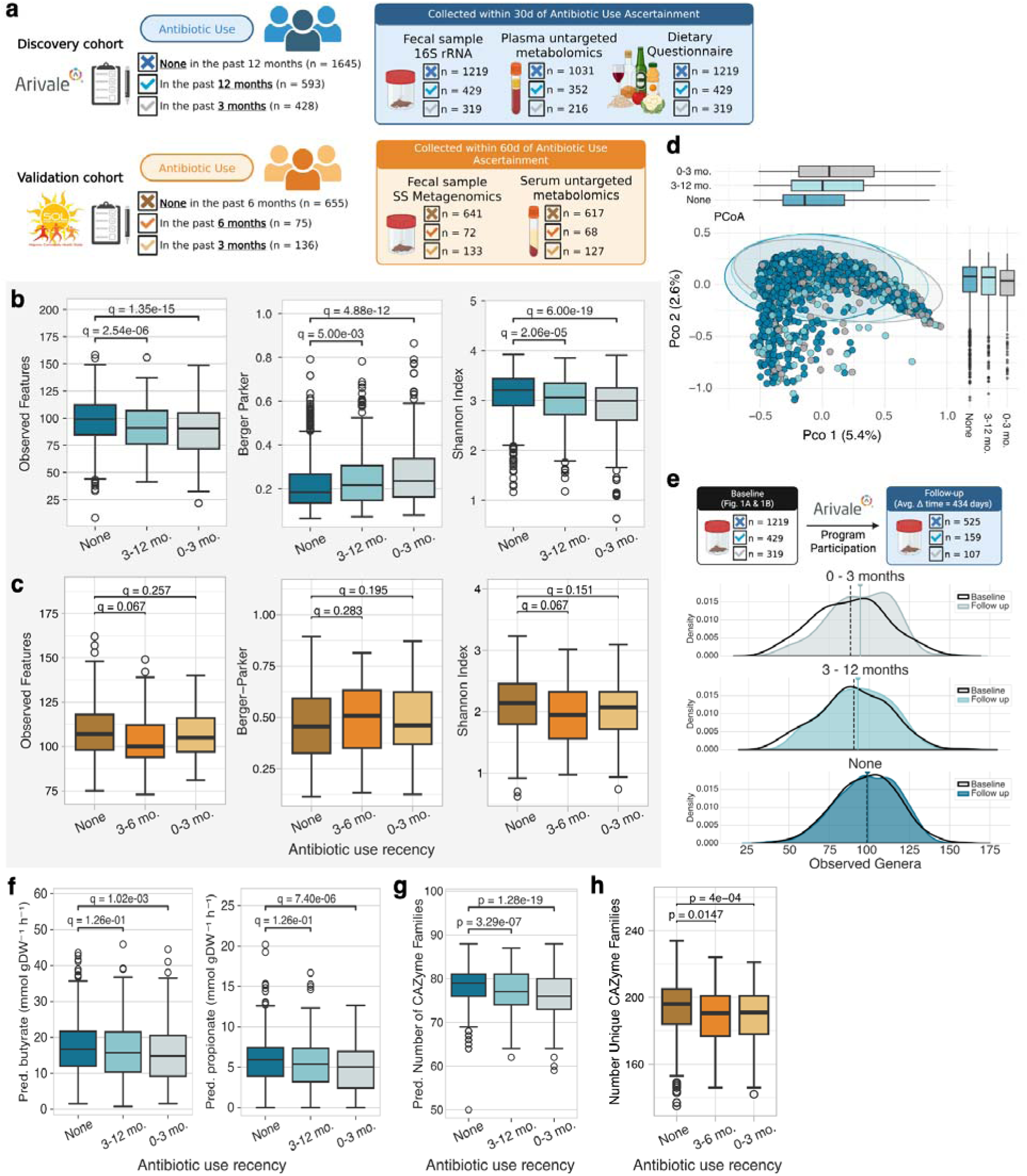
Characterizing gut microbiome community structure in individuals reporting recent antibiotic use. a, Antibiotic recency windows and distribution of biosamples available for each cohort. Answer choices are mutually exclusive and shown as presented to Arivale participants. **b,** Boxplots depicting alpha diversity metrics for Arivale: richness (left), dominance (center), and Shannon entropy (right). **c,** Alpha diversity metrics for HCHS/SOL are presented as in **b**. **d,** Principal coordinate analysis (PCoA) based on Bray-Curtis dissimilarity. Boxplots of antibiotic recency groups are presented on the side. Statistical significance was assessed using PERMANOVA with 9,999 permutations. **e,** Kernel Density Estimate (KDE) plots of observed genera for baseline and follow-up samples across the different antibiotic recency groups in Arivale. Black KDE plots show baseline data (presented in B), where the dashed black line represents the mean. Colored KDE plots show data for available follow-up stool samples, and solid-colored lines represent the mean for each group. **f,** Microbial community metabolic model (MCMM) predicted production of butyrate (left) and propionate (right). **g,** Boxplots showing the predicted number of CAZyme families generated with picrust2. **h,** Boxplots showing the number of CAZyme families estimated from metagenomic data with Cayman. Boxplots show median and interquartile range (IQR). Whiskers represent ±1.5×IQR, and points are outliers. Plots are of the unadjusted values while statistical annotations are derived from multivariable covariate-adjusted linear regression models with FDR correction using the BH method.

We used multivariable linear regression to assess cross-sectional variation in host molecular and microbiome features associated with recent antibiotic use. Models for the discovery cohort (Arivale) were adjusted for age, sex, BMI, estimated glomerular filtration rate (eGFR), the first five principal components of genetic ancestry, and cholesterol-lowering and blood sugar-lowering medication use (“full model”). Microbiome or clinical chemistry vendors were added as covariates, where applicable. For HCHS/SOL models, covariates included age, sex, study center, BMI, and eGFR. Birthplace was an additional covariate in microbiome models only. MC models were adjusted for gender, age, BMI, microbial load, patient group, study center, antilipid medication use, and antidiabetic medication use.

### Recent antibiotic use is associated with changes in the structure and function of the gut microbiome

Using linear regression models, we tested the association of antibiotic recency with three alpha diversity measures representing richness (observed features), dominance (Berger-Parker index), and a hybrid metric that takes both richness and evenness (inverse dominance) measures into account (Shannon index) ^47^. Both the 3-month and 12-month groups exhibited significantly fewer observed features and lower Shannon diversity than those without antibiotic use (two-tailed t-test, Benjamini-Hochberg [BH] q < 0.1). In contrast, we observed a significant positive relationship between the Berger-Parker index and the recency of antibiotic use (β_3_ = 0.05, BH q = 4.88 x 10^-12^; β_12_ = 0.02 BH q = 0.005), indicating that more recent antibiotic use is associated with decreased richness and increased dominance (Fig. 1B). In the HCHS/SOL cohort, estimates for all three metrics were directionally consistent with the Arivale findings, with reduced observed features and Shannon diversity in addition to increased Berger-Parker dominance in both recency groups relative to no reported use. However, in the HCHS/SOL cohort, only observed features in the six month group reached significance (β = −4.11, SE = 1.80, BH q = 0.07; Fig. 1C). Shannon diversity was similarly reduced in both the 3-(β = −0.09, SE = 0.05, BH q = 0.15) and 6-month groups (β = −0.11, SE = 0.06, BH q = 0.10).

Next, to examine how antibiotic use may influence interindividual variation in the gut microbiome, we performed principal coordinate analysis using a Bray-Curtis dissimilarity matrix computed from a rarefied ASV-level abundance matrix (21,869 reads per sample) in the Arivale cohort (Fig. 1D). PERMANOVA and PERMDISP revealed that the community structure and dispersion across the three antibiotic recency groups was significantly different (p-value = 0.0001; 9,999 permutations for each analysis). Follow-up pairwise comparisons confirmed that each group was significantly different from the others (BH q < 0.1; 9,999 permutations for all comparisons).

For a subset of 791 individuals in the Arivale cohort, a second stool sample was available at least 60 days after the first stool sample (mean time between samples: 434 days). Antibiotic recency was defined relative to the first stool sample (Fig. 1A & 1B) and held fixed for both timepoints; it describes how recently antibiotics had been used at baseline, not continued use between samples. We assessed the longitudinal change in gut microbiome alpha diversity using linear mixed-effects models (Fig. 1E). Using a likelihood ratio test (LRT) of models with and without the antibiotic recency and time interaction term, we determined that the trajectories of observed features and Shannon index differed by antibiotic recency group (BH q = 2.6 x 10^-4^ and 9.9 x 10^-3^, respectively). Estimated marginal slopes for observed features were monotonically ordered by recency (3 months: 1.33 genera per 100 days, 95% CI 0.70–1.96; 12 months: 0.46, −0.03–0.96; None: −0.08, −0.37–0.22), with the 3-month group differing from those reporting no recent use (Tukey-adjusted p = 1.0 x 10 ).

Having established that recent antibiotic use is associated with measurable impacts on alpha and beta diversity measures, we sought to investigate whether we could detect potential functional differences in the gut microbiome of recent antibiotic users. We used microbial community-scale metabolite models (MCMMs) ^48^ to predict short-chain fatty acid (SCFA) production in Arivale participants, using microbiome data and fiber intake estimates from dietary questionnaires as constraints ^49^. We tested the Arivale antibiotic recency groups for differences in predicted butyrate and propionate production (Fig. 1F), finding that the gut microbiomes of individuals in the 3-month group were predicted to produce less butyrate (β = −1.46, BH q = 0.001) and propionate (β = −0.81, BH q = 7.4 x 10^-6^).

Given that we were working with 16S amplicon sequencing data in the Arivale cohort, we used Picrust2 ^50,51^ to generate carbohydrate-active enzyme (CAZyme) family gene abundance predictions (Fig. 1G), finding that both the 3- and 12-month groups harbor fewer predicted CAZymes (two-tailed t-test, p < 0.05). Moreover, we leveraged HCHS/SOL stool shotgun metagenomic data to further investigate the CAZyme repertoire following antibiotic use. In these samples, CAZyme annotations were generated using Cayman ^52^. Fewer unique CAZyme families were observed in both 3- and the 6-month groups, compared to the group reporting no antibiotic use within the past 6 months (Fig. 1H; two-tailed t-test, p < 0.05), and the overall structure of the CAZyme repertoire was significantly different between the antibiotic recency groups (Fig. S2, PERMANOVA, R^2^ =0.004, p = 0.002).

Finally, we observed an enrichment of *Bacteroides*-dominated enterotypes in antibiotic users across both Arivale and HCHS/SOL cohorts (Fig. S3). In Arivale, the strongest enrichment was for the Bac2 enterotype (one of the four enterotypes assigned to Arivale participants, using a Dirichlet Multinomial Mixture model ^53^ , which was enriched in the genus *Bacteroides* and depleted in butyrate producers) in individuals reporting antibiotic use in the last 3 months (OR = 2.89, BH q = 6.96 x 10^-13^, two-sided Wald test) and in the last 12 months (OR = 1.75, BH q = 4.04 x 10^-05^, two-sided Wald test). This was accompanied by a concomitant decrease in the proportion of individuals assigned to the Ruminococcaceae (Rum) enterotype in recent antibiotic users (OR_3_ = 0.48, BH q = 1.23 x 10^-06^; OR_12_ = 0.54, BH q = 9.62 x 10^-06^, two-sided Wald tests).

### Genus-level associations with antibiotic recency

Having detected high-level structural differences in the gut microbiome of recent antibiotic users, we next investigated variation in gut microbiome composition using two approaches. First, we investigated potential changes in centered log ratio (CLR) transformed relative abundances (Fig. 2, inner heatmap) by constructing individual linear regression models for all genera with prevalence above 10% in the Arivale cohort (n = 188, Table S2). In line with previous studies of antibiotic use in human cohorts^18,54^, we observed increased abundances of *Enterococcus*, *Veillonella*, and *Escherichia/Shigella* in individuals who reported recent antibiotic use (BH q < 0.1). However, many of these findings were significant only in the 3-month group, indicating that antibiotic-associated differences in the abundances of gut bacterial genera were dampened over longer recovery times. Additionally, we observed an increase in the relative abundance of *Sellimonas* (β_3_ = 0.73, two-tailed t-test, BH q = 5.46 x 10^-08^), which has recently been proposed as a marker of post-disturbance recovery in the gut ^55^. Because individuals with recent antibiotic use have decreased gut microbiome richness, some genera may be below the limit of detection or have failed to recover in these individuals. Thus, we also used logistic regression models to identify genera (n = 209, Table S3) with higher or lower odds of being detected in the gut microbiome of antibiotic users (Fig. 2, outer heatmap). Most genera were associated with decreased odds of detection in the gut microbiome of recent antibiotic users (OR < 1), consistent with reduced richness following antibiotic exposure. A minority, however, showed the opposite trend: 17 genera had increased odds of detection in at least one of the two recent antibiotic use groups, predominantly low-prevalence genera. *Citrobacter*, for instance, was detected in 21/319 (6.6%) of participants reporting antibiotic use within the last three months and 25/429 (5.8%) within twelve months, compared to 27/1219 (2.2%) of non-users (OR_3_ = 2.89, BH q = 0.001; OR_12_ = 2.58, BH q = 0.006).

**Figure 2.**
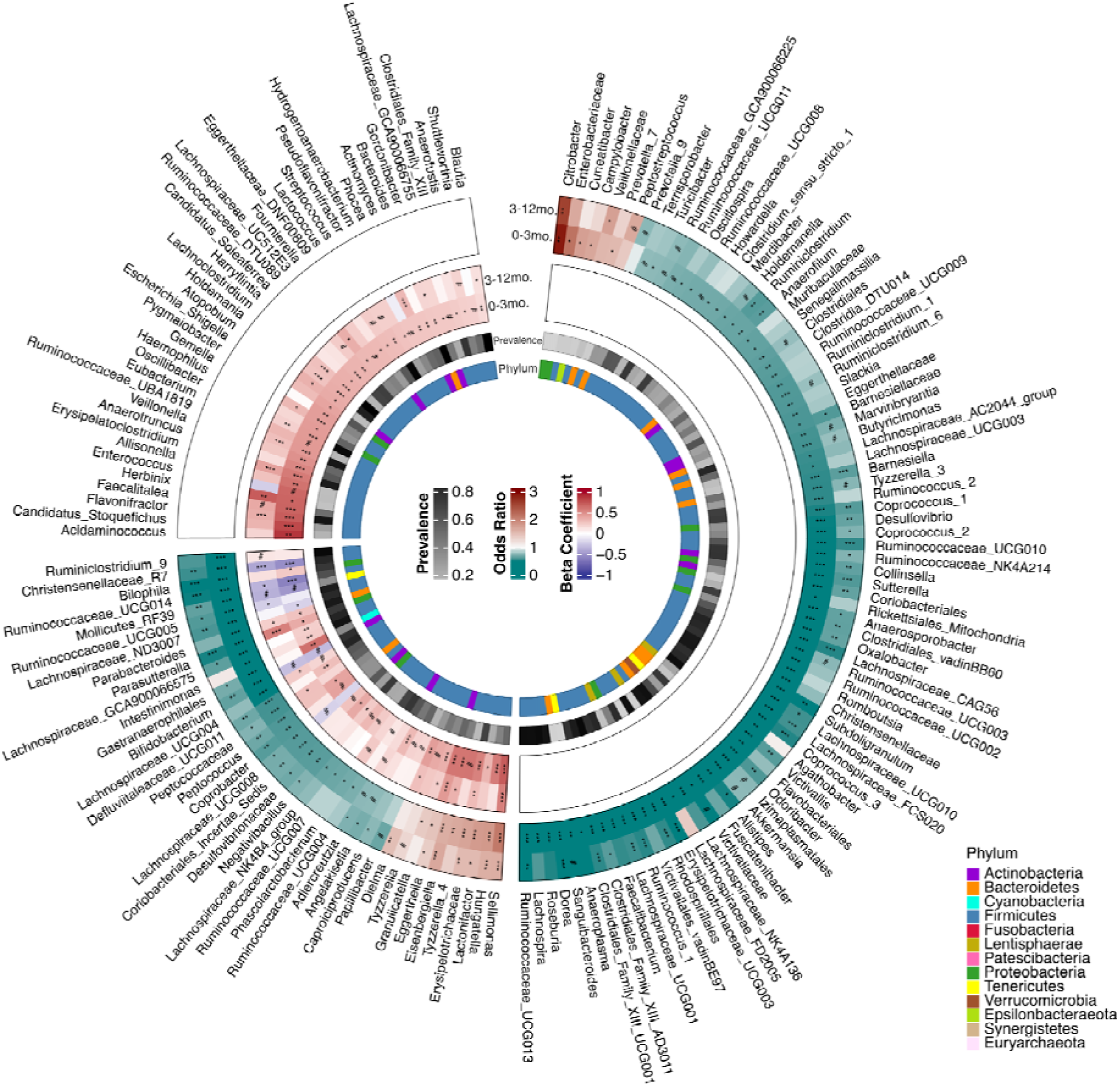
Antibiotics are associated with changes in relative abundance and presence/absence. Circular heatmaps depicting beta coefficients for taxon CLR abundance (inner track) and logistic regression odds ratios for taxon presence/absence (outer track). Arcs within tracks represent values for the 3-month (inner) and 12-month (outer) groups. The number of taxa tested in logistic (n=209) and linear regressions (n=188) differed because of different prevalence filtering thresholds. Sectors show taxa significant in only linear regression models (top left), shared hits (bottom left), and taxa significant in only logistic regression models (right). Linear models were adjusted for all covariates in the “full model”. FDR correction was performed using the BH method: # < 0.1, * < 0.05, ** < 0.01, *** < 0.001.

For a small subset of genera, there were inconsistent trends between the logistic and linear regressions. Certain genera were associated with increased odds of detection in recent antibiotic users and, when present, were more abundant than in non-users—like *Sellimonas,* for example. However, genera such as *Gastranaerophilales* and *Parasutterella* had lower odds of being detected among recent antibiotic users but then showed an increase in relative abundance in antibiotic users when they were present (Fig. 2).

### Recent antibiotic use impacts the blood metabolome, but not the blood proteome

We next investigated whether or not we could detect variation in the blood metabolome and proteome associated with recent antibiotic use. Starting with the Arivale cohort, we built individual linear regression models for metabolites with over 70% prevalence (n = 969, Table S4), finding 32 metabolites with altered abundance in the 3-month group (Fig. 3A, two-tailed t-tests, BH q < 0.1). Only 5 were altered in the 12-month group, reflecting similar recovery trends observed in the gut microbiome diversity and composition. Two unannotated metabolites (X-12283 and X-17351) were significantly different in the 12-month group but not in the 3-month group, suggesting a temporal association or unmeasured group-specific differences. We refer to the union of these two sets (n = 34 metabolites) as antibiotic-responsive metabolites. In most cases, metabolites were significantly reduced in abundance in recent antibiotic users compared with the non-users. These antibiotic-responsive metabolites include host-microbe co-metabolites, and the majority were shown to be more closely associated with the gut microbiome than with host genetics in a prior study ^30^. Finally, we assessed whether recent antibiotic use is associated with differences in the blood proteome. After applying a 70% prevalence threshold, we tested 274 proteins (adjusting for the full set of covariates), finding no significant differences in the levels of these circulating proteins between antibiotic users and non-users (Table S5).

**Figure 3.**
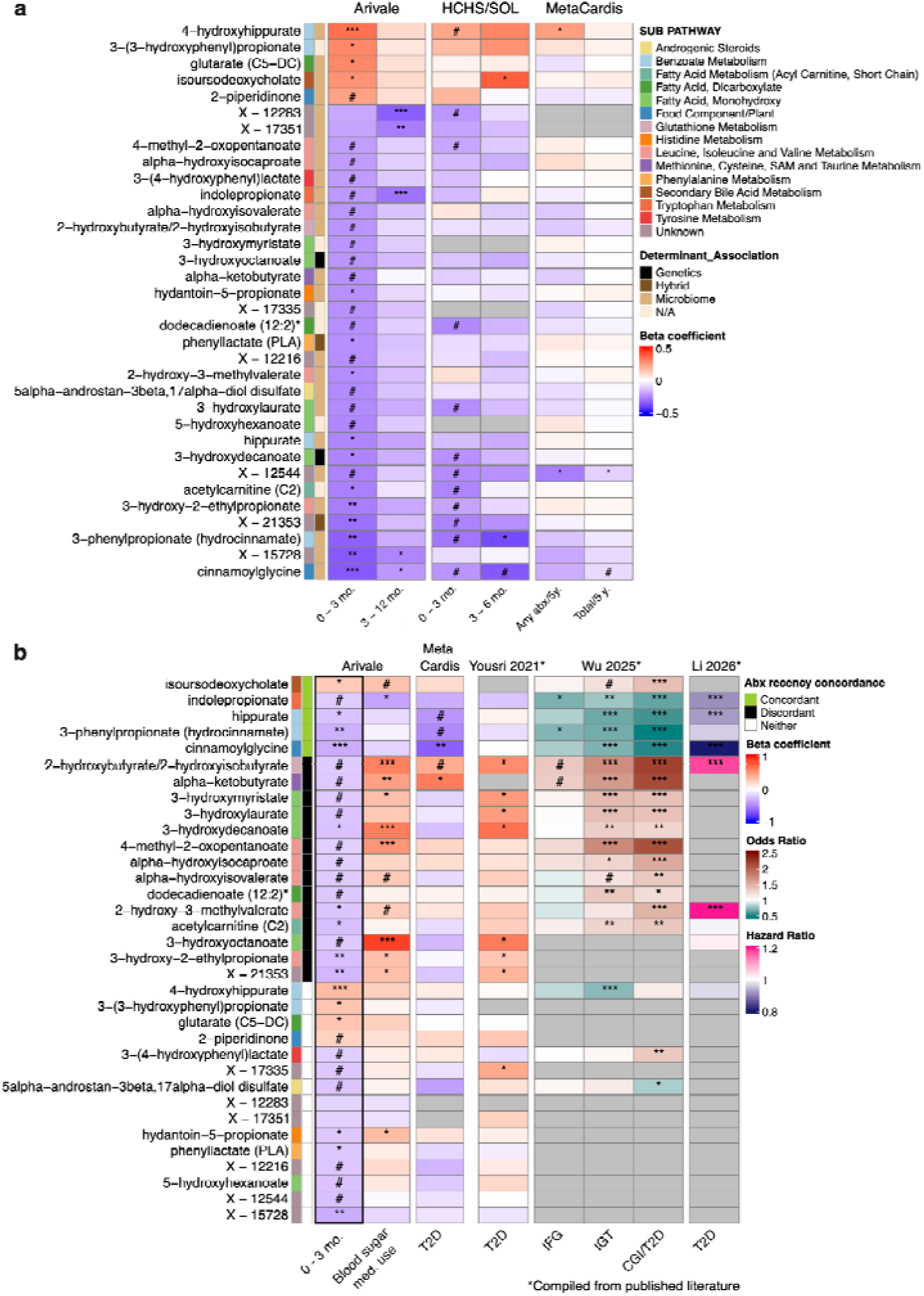
Antibiotics are associated with changes in 34 blood metabolites, several of which are associated with glucose intolerance phenotypes. a, Heatmaps of linear regression coefficients for antibiotic use in Arivale (left), HCHS/SOL (middle), and MetaCardis (right). Columns for Arivale and HCHS/SOL represent antibiotic recency windows: 0-3 months (left), 3-12 months (right, Arivale), or 3-6 months (right, SOL). MetaCardis columns: binarized antibiotic use (left) and total number of courses in the previous five years (right). FDR correction was performed using the BH method: # < 0.1, * < 0.05, ** < 0.01, *** < 0.001. Determinant annotations were derived from Diener et al. ^30^ **b,** Heatmap displaying the associations of antibiotic-responsive metabolites with T2D phenotypes. Coefficients for the 3-month antibiotic use group are shown as a reference (left; black border). Associations for blood sugar-lowering medication (Arivale, second from left) and T2D (MetaCardis, third from left) are shown next. The remaining columns show results from publicly available supplementary data (denoted with *) from Yousri et al., ^59^ Wu et al., ^60^ and Li et al. ^61^ For these datasets, estimates and adjusted p-values are displayed as provided in the source (beta coefficient (Bonferroni), OR (BH FDR), and HR (BH FDR), respectively). Antibiotic recency concordance was determined by whether a metabolite showed a consistent association with glucose intolerance phenotypes in two patient groups or cohorts: green (concordant), black (discordant), neither (white). IFG: impaired fasting glucose, IGT: impaired glucose tolerance, CGI: combined glucose intolerance.

To assess the robustness of the metabolite associations, we next pursued validation in two external cohorts, constructing individual linear regression models for only the 34 antibiotic-responsive metabolites, if present in the external cohort. In the HCHS/SOL cohort, 13 of these metabolites, including several amino acid and fatty acid derivatives, were statistically significantly different in the recent antibiotic users, and the directionality was consistent (BH q < 0.1). We next validated these results in the MC cohort ^1^. Because antibiotic use in the previous 5 years is enumerated in MC, we used two approaches for validation in this cohort: binarizing the variable to assess the impact of using any number of courses over the previous 5 years (most similar to Arivale), and using the total number of courses as a continuous variable. Unsurprisingly, given the longer antibiotic recency time window, fewer metabolites were statistically significantly altered in the MC cohort (N=3, BH q < 0.1). In particular, differences in cinnamoylglycine, 4-hydroxyhippurate, and X-12544 levels were statistically significant and directionally consistent among antibiotic users in all 3 cohorts.

#### Antibiotic-responsive blood metabolites show variable associations with metabolic health status

We next assessed the association between the 34 antibiotic-responsive metabolites and T2D phenotypes (Fig. 3B, Table S6). Blood sugar-lowering medication use was included as a covariate in the Arivale models, as medication intake can account for a substantial fraction of apparent disease-metabolome associations ^1^. Examining this coefficient, 12 of the 34 antibiotic-responsive metabolites were higher in Arivale participants reporting blood sugar-lowering medication use (including branched-chain amino acids catabolites and monohydroxy fatty acids), while indolepropionate was lower (BH q < 0.1). Because MC includes individuals along the cardiometabolic spectrum, we also examined the patient group coefficient contrasting T2D against healthy individuals, adjusted for antidiabetic medication use. Individuals with T2D in MC had lower levels of hippurate, 3-phenylpropionate, and cinnamoylglycine, consistent with prior reports associating lower hippurate with metabolic syndrome ^56,57^ and of reduced 3-phenylpropionate and cinnamoylglycine with T2D ^58^.

We next compiled results from three published studies investigating the relationship between circulating metabolites and glycemic phenotypes in diverse cohorts ^59–61^. Yousri et al. built linear regression models for 826 metabolites and prevalent T2D in a Qatari cohort, validating the results with data from the Qatar Biobank ^59^. Wu et al. pursued serum metabolite associations with impaired fasting glucose, impaired glucose tolerance, and combined glucose intolerance/T2D in two independent Swedish cohorts ^60^. Finally, Li et al. examined 469 metabolites in relation to incident T2D across ten population-based cohorts (including SOL) of initially T2D-free participants followed for up to 26 years ^61^. Thus, effect estimates are not comparable (betas [Yousri et al.], odds ratios [Wu et al.], and hazard ratios [Li et al.], respectively; Fig. 3B), but the directions of association can be compared across these designs. Of the three studies, the linear regression models built by Yousri and colleagues most closely resemble the Arivale models, and the direction of association between the antibiotic-responsive metabolites and prevalent T2D is also largely consistent with blood sugar medication use associations in Arivale. Findings from the latter two studies are sparse because the regressions were limited to results observed in both discovery and validation cohorts (Wu et al.) or metabolites successfully harmonized across the 10 cohorts (Li et al.). Notably, compiling these prior associations lets us visualize divergence between antibiotic-responsive and glucose intolerance-associated metabolites. That is, for five metabolites—isoursodeoxycholate, indolepropionate, hippurate, 3-phenylpropionate, and cinnamoylglycine—the direction of association with phenotypes along the glucose-dysregulation spectrum is concordant with recent antibiotic use across at least 2 patient groups or studies. Meanwhile, 14 metabolites, such as alpha-ketobutyrate and 4-methyl-2-oxopentanoate, were discordant: depleted in recent antibiotic users yet positively associated with glucose dysregulation. The remaining 15 metabolites had no clear association with glucose dysregulation phenotypes. Metabolites positively associated with metabolic health, such as cinnamoylglycine ^58^, hippurate ^56^, and indolepropionate ^62^, were depleted in recent antibiotic users. However, branched-chain amino acid catabolites—which have been positively associated with T2D ^63^—were also decreased in antibiotic users, suggesting heterogeneous or context-dependent associations of antibiotic exposure on microbial metabolites relevant to host metabolic homeostasis.

### Moderation effects by diet and gut microbiome composition reveal context dependencies

To explore context-dependent associations with recent antibiotic use, we investigated diet as a potential moderator of the effect of antibiotics on gut microbiome diversity in the Arivale cohort (Fig. 4A, Table S7). Arivale participants completed health questionnaires, including limited food frequency questionnaires, which assessed habitual diet. Most dietary items were sparsely populated in our cohort, and additional variables were excluded during preprocessing, leaving 9 dietary variables for analysis (Methods; Table S7). Focusing on these variables, we binarized all dietary features that capture intake frequency to balance group sizes across categories, and we tested the three alpha diversity metrics presented above (Fig. 1) for moderation effects. Moderation effects were only found for the number of observed features, where a preference for liquor (Fig. 4B) or an intake of at least one alcoholic drink per day (Fig. 4C) was associated with lower gut microbiome richness in the 3-and 12-month groups, respectively (interaction term BH q < 0.1, two-tailed t-test). The 3-month group also displayed interesting trends for daily intake of sugary drinks and fruit: consumption of at least one serving of sugary drink per day may be associated with lower gut microbiome richness, whereas consumption of at least one serving of fruit per day may be associated with increased gut microbiome richness (Fig. 4C, 0.1 < BH q < 0.2).

**Figure 4.**
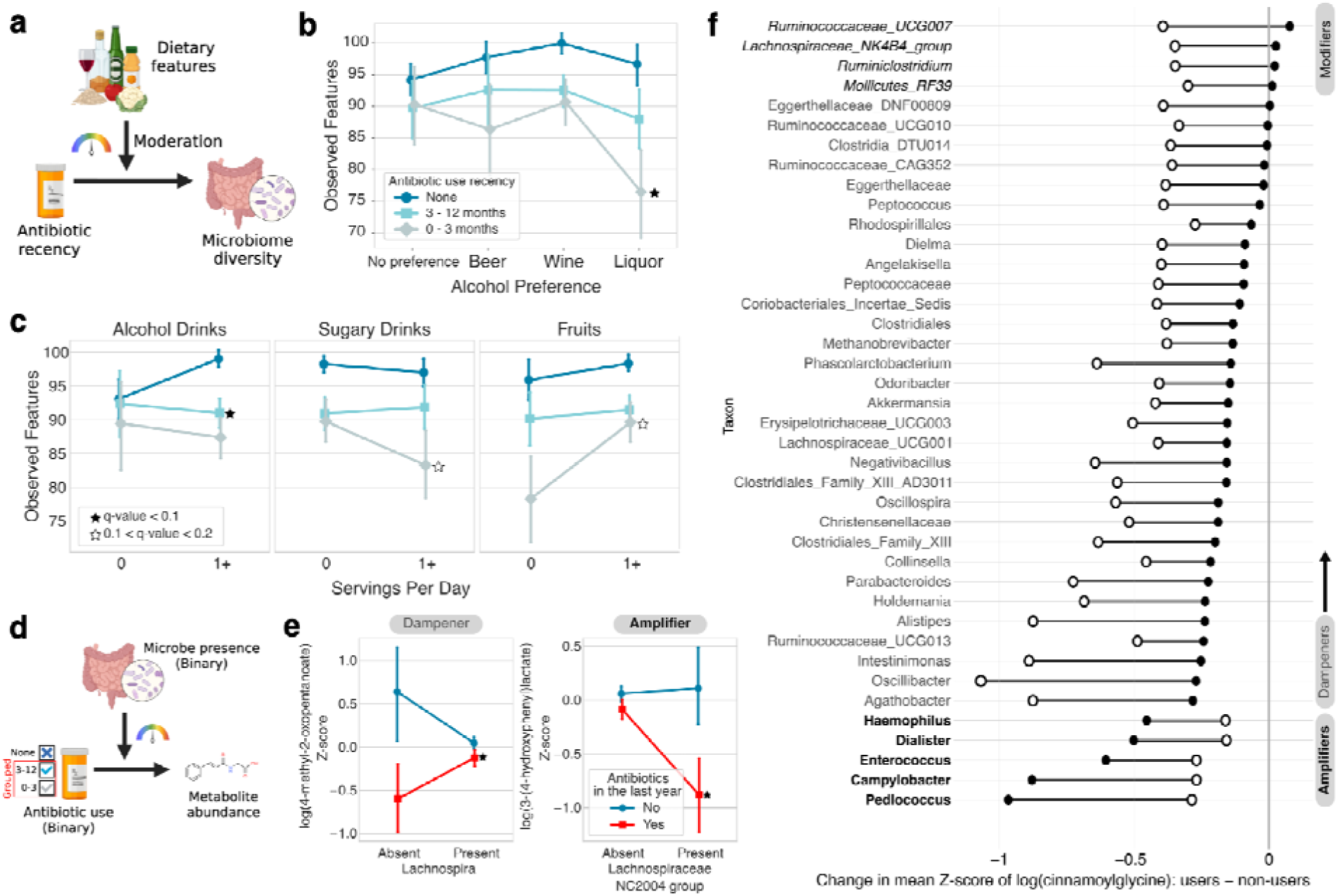
Moderation by diet and microbiome composition a, Schematic of dietary variables moderating the effect of antibiotics on alpha diversity **b,** Point plot demonstrating the interaction between alcohol preference and antibiotic recency. **c,** Point plots demonstrating the interaction between binarized dietary variables and antibiotic recency. **d,** Schematic of binarized microbe presence/absence moderating the impact of antibiotic use (binarized) on metabolite abundance. **e,** Point plots displaying the difference in normalized blood 4-methyl-2-oxopentanoate (left) and 3-(4-hydroxyphenyl)lactate abundance in antibiotic users (red) and non-users (blue). **f,** Barbell plot showing the difference in mean normalized blood cinnamoylglycine levels between antibiotic users and non-users (interaction term BH q <0.1 for all genera shown). Closed and open circles represent genus presence or absence, respectively. Genera considered “amplifiers” are denoted in bold, “modifiers” are italicized in black, and “dampeners” are in grey normal typeface. For point plots, stars indicate an interaction effect with BH q-value thresholds of 0.1 (closed) or 0.2 (open), respectively.

Next, we wondered whether gut microbiome composition could moderate the effect of antibiotic treatment on the blood metabolome. For these regressions, we used binarized microbial presence or absence (n = 209 taxa, Table S8) as the moderator and built individual moderation models for each antibiotic-responsive metabolite and microbe pair (n = 7106 models; Fig. 4D). We also binarized antibiotic use recency due to the sparsity of these data types (e.g., for some microbe-metabolite pairs, there were no observations in one of the 3- or 12-month groups). We found moderation effects for ten metabolites: cinnamoylglycine, 3-phenylpropionate (hydrocinnamate), hippurate, 2-piperidinone, glutarate (C5-DC), hydantoin-5-propionate, 3-(3-hydroxyphenyl)propionate, 3-(4-hydroxyphenyl)lactate, 4-methyl-2-oxopentanoate, and the unannotated metabolite X-12544 (interaction term BH q-value < 0.1, two-tailed t-tests). Some metabolites possessed few microbial moderators (Fig. 4E, Fig. S4). *Lachnospira*, for instance, was associated with a smaller decrease in 4-methyl-2-oxopentanoate levels in those reporting antibiotic use in the last year, whereas the presence of *Lachnospiraceae NC2004 group* was associated with a larger decrease in 3-(3-hydroxyphenyl)propionate in the antibiotic-treated group. In contrast, we identified over five potential microbial moderators for cinnamoylglycine, 3-phenylpropionate (hydrocinnamate), hippurate, 2-piperidinone, and glutarate (C5-DC). In most cases, microbial moderators reduced the effect of recent antibiotic use on the blood metabolome (i.e., ‘dampeners’; Fig. 4F). We observed instances where the antibiotic-metabolite association reverses direction in the presence of a particular genus (i.e., ‘modifiers’; Fig. 5F and Fig S4). Finally, the presence of certain genera, such as *Dialister* and *Veillonella*, was associated with a greater change in blood metabolite abundance in recent antibiotic users compared to non-users (i.e., ‘amplifiers’).

**Figure 5.**
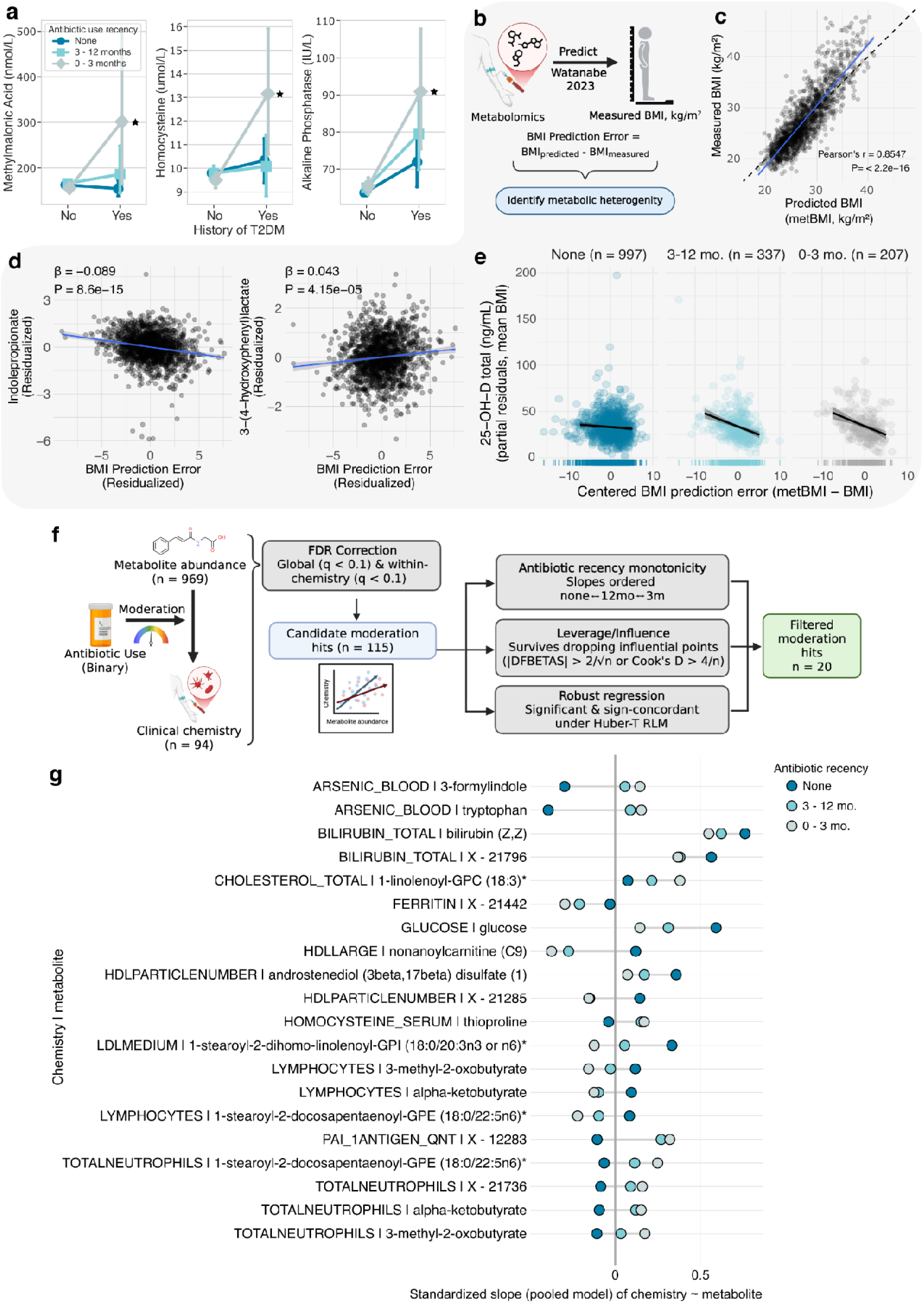
Recent antibiotic use is associated with variation in clinical chemistries, but only in certain metabolic health contexts. a, Point plot demonstrating the interaction between history of T2D (none: n = 42, 12-month: n = 24, and 3-month: n = 18) and recent antibiotic use on clinical chemistries. Black stars indicate a significant interaction effect (FDR q-value < 0.1). **b,** Schematic illustrating the metabolic BMI (metBMI) machine learning model devised by Watanabe et al., and our approach for leveraging BMI prediction error to identify metabolic heterogeneity. **c,** Pearson correlation between measured and predicted BMI (r = 0.85, P < 2.2e-16). **d,** Association between BMI prediction error and blood indolepropionate (left) and 3-(4-hydroxyphenyl)lactate levels (right). P-values were calculated with two-sided t-tests. **e,** Association between total 25-Hydroxyvitamin D levels and the interaction between antibiotic recency and BMI prediction error. **f,** Schematic describing moderation analysis focusing on metabolite abundances’ effect on clinical chemistries. Hits were subject to within-chemistry and global significance thresholds (FDR < 0.1) and underwent the following filtering steps: checking antibiotic recency monotonicity, surviving dropping influential points, and robust regression. **g,** Barbell plot showing the 20 significant associations moderated by antibiotic use. Standardized slopes for each antibiotic recency group are shown in dark blue (none), teal (12-month), and grey (3-month).

### Context-dependent associations between recent antibiotic use and clinical chemistries

Prior studies investigating the effect of antibiotic use on long-term metabolic health outcomes have produced mixed results ^23^. To explore this inconsistency in the Arivale cohort, we next constructed covariate-adjusted linear regression models for 94 clinical chemistry outcomes. We found no significant differences in clinical chemistries between recent antibiotic users and non-users (Table S9), consistent with prior results from smaller placebo-controlled randomized trials ^18,19^. We next performed moderation analyses to explore potential subpopulation heterogeneity. First, we constructed linear regression models for all 94 chemistries as done previously, but adding a history of T2D (none: n = 42, 12-month: n = 24, and 3-month: n = 18) as a moderator (Table S9). We found significantly higher levels of methylmalonic acid, homocysteine, and alkaline phosphatase in individuals with T2D having antibiotic use in the last three months (Fig. 5A, two-tailed t-test, BH q < 0.1).

Given the variable associations of individuals with a history of T2D, we next turned to previous work in the Arivale cohort that established metabolomics-inferred measures of BMI (metBMI) as more sensitive to lifestyle changes and metabolic heterogeneity ^64^. We wondered whether we could use metBMI to detect subtle differences in host metabolic health following antibiotic use. Recent antibiotic users were excluded from the original metBMI study, so we applied the beta coefficients from their metBMI model to generate new metBMI predictions for antibiotic users in the Arivale cohort. To assess metabolic heterogeneity, we calculated metBMI error as the difference between predicted (metBMI) and measured BMI, and applied an interaction with the antibiotic recency variable (Table S10). We tested 94 clinical chemistries and found two that passed our significance threshold (Type II ANOVA, interaction term; BH q < 0.1): absolute basophils and 25-hydroxyvitamin D, 25(OH)D. The association between absolute basophils and metBMI error diverged between those with antibiotic use in the previous three months (positive association) versus use in the prior twelve months (negative association). For 25(OH)D, the metBMI-moderated antibiotic recency coefficients were negative and monotonic. In other words, as metBMI error increases—suggesting worse metabolic health than expected using measured BMI alone—25(OH)D levels decrease more strongly among more recent antibiotic users. This association was robust to further adjustment for vitamin D supplementation and season, and removing two outliers with levels of 25(OH)D greater than 150 mg/dL.

Finally, we performed a large, unbiased screen of antibiotic-moderated associations between blood metabolite abundances (n = 969) and clinical chemistry outcomes (n = 94). Here, our goal was to learn more about how recent antibiotic use (binarized) might change the relationship between blood metabolites and clinical chemistries (Fig. 5F, Table S11). After applying a global (BH q < 0.1) and within-family (BH q < 0.1) FDR correction, we filtered our results by requiring hits to exhibit monotonic antibiotic recency trends, survive dropping influential points, and remain significant with robust regression. We found 20 associations where antibiotic use moderated the effect of metabolite abundance on clinical chemistries (Fig. 5G). Encouragingly, we observed a positive association between features detected in both the metabolomics and clinical chemistry panels (e.g., glucose and bilirubin). Except for the association between LDL medium particles and 1-stearoyl-2-dihomo-linolenoyl-GPI (18:0/20:3n3 or n6)*), moderation by recent antibiotic use exhibited two patterns. Most often, the metabolite-clinical chemistry association exhibited opposing directions between recent antibiotic users and non-users. For example, the association between the metabolites tryptophan and 3-formylindole with blood arsenic levels is negative in those without recent antibiotic use, whereas this association is positive in recent antibiotic users. Further, in recent antibiotic users, abundances of alpha-ketobutyrate, 3-methyl-2-oxobutyrate, and 1-stearoyl-2-docosapentaenoyl-GPE (18:0/22:5n6)* were positively associated with neutrophil percentage, whereas the association was negative in non-users. We see the opposite pattern for lymphocyte percentage, which is unsurprising given that neutrophil and lymphocyte percentages are compositional measures normalized to the total number of white blood cells. Incidentally, moderation of absolute neutrophil or lymphocyte counts did not reach statistical significance, though the coefficients were directionally consistent. Finally, in select cases, the metabolite-chemistry association was directionally consistent, though the magnitude of the beta coefficient varied in recent antibiotic users. We observed this pattern in related molecules, such as the association between bilirubin and X-21796, which was recently hypothesized to be a bilirubin derivative ^65^.

## Discussion

Contrary to epidemiological evidence linking antibiotics to heightened risk of T2D ^6,8^ , short-term intervention studies have not been able to link antibiotic use to long-term changes in metabolic parameters or hormones, despite showing that antibiotics can alter gut microbiome composition ^18,39,54^. Here, we leverage three cohorts, encompassing diverse participant demographics, to assess the molecular signatures of recent antibiotic use. We first investigated differences in gut microbiome diversity, function, and community composition associated with recent antibiotic users. As expected, recent antibiotic treatment is associated with decreased gut microbiome richness (Fig. 1B) and altered community structure (Fig. 1D). In Arivale, we observed a clear monotonic relationship between the recency of antibiotic treatment and the magnitude of the association with gut microbiome diversity and composition, suggesting recovery of the gut microbiome over time. Accordingly, in our longitudinal analysis of gut microbiome richness, individuals who had used antibiotics in the 3 months prior to their baseline stool sample experienced a greater positive change in the number of observed genera over time than those reporting no antibiotic use in the past year, with the 12-month group showing an intermediate slope (Fig. 1E). This graded pattern is consistent with the broader observation that antibiotic-associated diversity loss is generally followed by rapid recovery within a few weeks of treatment ^2,66^. Furthermore, we found that enterotypes dominated by *Bacteroides* were enriched in recent antibiotic users, while the enterotype associated with higher levels of butyrate-producing Clostridia was depleted (Fig. S3). Antibiotic enrichment for *Bacteroides* dominance may have negative implications in the context of aging, given that this genus was previously associated with poorer metabolic side effects in response to statins and with reduced survival in the latest decades of life ^67,68^. We also observed expected functional differences in the gut microbiome of recent antibiotic users, including lower predicted SCFA production (Fig. 1F) and decreased CAZyme richness (Fig. 1G & 1H). Though our analyses were cross-sectional, these antibiotic recency groups may represent windows along a trajectory of gut microbiome recovery. The monotonic trends in CAZyme richness seen in both Arivale and HCHS/SOL were consistent with previous work by Chng et al.^69^, suggesting that the number of carbohydrate-active enzyme (CAZyme) families is positively correlated with community recovery from antibiotic perturbations.

We also investigated individual genus-level differences in the gut microbiome of recent antibiotic users (Fig. 2). Genera containing opportunistic enteric pathogens (e.g., *Citrobacter* and *Campylobacter*) showed increased odds of detection, consistent with prior studies^1,2^, and we observed increased relative abundance of oral-associated bacteria, such as *Veillonella*. *Tyzzerella 4* was increased in both detection and relative abundance in individuals reporting antibiotic use in the previous 3 months relative to non-users, in line with a previous study associating this taxon with antibiotic use and adiposity in children ^70^. *Tyzzerella 4* may also mediate the effect of smoking on risk of colorectal high-risk adenoma ^71^, highlighting the interconnectedness of host behavior and gut microbiome effectors. For some antibiotic-responsive taxa, such as *Parasutterella* and *Gastranaerophilales*, we found lower odds of detection in recent antibiotic users, but an increased relative abundance in antibiotic users, relative to non-users, when the taxon was detected. This phenomenon may arise due to stochastic extinction of low-abundance taxa (such as *Gastranaerophilales*) during antibiotic-induced population bottlenecks, which are then able to bloom in the antibiotic-disrupted ecosystem if they avoid local extinction.

During our investigation of the blood metabolome following antibiotic treatment, we found 34 metabolites, including several fatty acid and amino acid metabolites, that were significantly different between recent antibiotic users and non-users (Fig. 3A). For nearly all of these metabolites, prior work found that the variance in these molecules was explained solely by the microbiome and not host genetics ^30^. However, we saw that 3-hydroxyoctanoic and 3-hydroxydecanoic acids were depleted in antibiotic users, even though these metabolites were previously only found to be associated with host genetics in the Arivale cohort ^30^. This result is in line with prior work showing that 3-hydroxyoctanoic acid is produced by *Parabacteroides goldsteinii* in culture ^72^, 3-hydroxydecanoic acid is elevated in the bloodstream of germ-free mice ^73^, and adds to a growing body of evidence that antibiotic treatment is a useful tool for identifying microbiome-responsive metabolites ^34^. Moreover, the majority of antibiotic-responsive metabolites decreased in abundance, in line with antibiotic-mediated disruption of bacterial transformations. For instance, decreased levels of 3-phenylpropionate, hippurate, and cinnamoylglycine could be the result of decreased bacterial phenylalanine metabolism ^74–76^, leading to lower levels of microbe-host co-metabolism ^36,77^. Meanwhile, we detected increased levels of 3-(3-hydroxyphenyl)propionate, along with a concomitant increase in the relative abundance of flavone metabolizing genera, *Flavonifactor* and *Eggerthella* ^78^. These antibiotic-associated differences in the host metabolome may have variable associations with health and physiology (Fig. 3B). For example, several BCAA metabolites, known biomarkers of metabolic diseases ^63^, were reduced in abundance following antibiotic use. However, isoursodeoxycholate, which has been associated with post-prandial lipemia and elevated liver enzymes in individuals recovering from bariatric surgery ^79^, was more abundant in those who had used antibiotics in the last three months. In contrast to the close association between the microbiome and the antibiotic-responsive metabolite hits, we detected no differences in the blood proteome following recent antibiotic use, consistent with prior work in the Arivale cohort indicating that the microbiome shows weaker coupling to the blood proteome than it does to the blood metabolome ^31^.

To investigate factors that may lead to variability in the physiological response to antibiotics, we looked at dietary behaviors and gut microbiome composition as potential moderators. We found that a preference for liquor was associated with lower gut microbiome richness in antibiotic users (Fig. 4B). Despite not meeting a significance threshold of q < 0.1, we observed a similar trend of lower richness in antibiotic users consuming at least one sugary drink per day and higher richness in individuals who consumed fruit (Fig. 4C), consistent with observational ^80,81^ and *in vitro* studies ^27^. Having detected dietary variables that influence microbiome recovery, we next moved on to investigating the influence of gut microbiome composition on host responses to antibiotic use. Chng et al. describe recovery-associated bacteria that are key to ecological recovery of the gut microbiome through successional processes, and that, importantly, the combined influence and abundance of these species is critical for recovery ^69^. In line with this observation, we found several significant genus-antibiotic interaction effects that help to explain variation in blood metabolite levels (Fig. 4D). Specifically, we found that microbial moderators could dampen or amplify antibiotic-associated differences in blood metabolite abundances (Fig. 4E). Commensal microorganisms known to produce beneficial metabolites, such as SCFAs, were often associated with dampening effects, whereas the opposite was true for orally-derived taxa and opportunistic pathobionts. Cinnamoylglycine had 40 candidate microbial moderators (Fig. 5F). This was the highest number of potential moderators detected, in line with cinnamoylglycine’s close association with alpha diversity metrics ^31,82^. Moreover, the presence of *Enterococcus,* previously observed to expand under sugary diets following antibiotic treatment ^80^, was associated with a greater loss in blood cinnamoylglycine levels (Fig. 5F), affirming the importance of assessing both host-behavior and microbiome variables.

We next investigated whether 94 clinical chemistry measures, including glycohemoglobin A1c, total cholesterol, and C-reactive protein, varied significantly between antibiotic users and non-users, adjusting for the full set of Arivale covariates (Table S9). On average, there were no covariate-adjusted associations between antibiotic use and clinical chemistries across the entire population, which aligns with results from intervention studies ^18,19^. Suspecting subpopulation heterogeneity, we used both medical history and a metabolomic-inferred BMI metric ^64^ to evaluate if antibiotic-associated perturbations of clinical chemistry measures differed by metabolic health status (Fig 5A-E). We found increased levels of methylmalonic acid and homocysteine in those with T2D who had taken antibiotics in the past three months—but not in the past 12 months—perhaps indicating a transient, antibiotic-associated vitamin B12 deficiency, but only in individuals with T2D ^83^. Vitamin B12 deficiency is a known side effect of metformin treatment ^84^, and microbial mechanisms have been hypothesized ^1,85^. Indeed, the gut microbiome may directly provision B12 to the host, although host absorption of B12 in the colon is thought to be limited ^86–89^. Given there were no average differences in clinical chemistries between antibiotic users and non-users in our original model, these findings could point to a short-term disruption of B12 balance that is only detectable in those at greater risk of deficiency. We also observed elevated alkaline phosphatase levels in the blood of recent antibiotic users with T2D, relative to non-users; however, this was not accompanied by a statistically significant difference in gamma-glutamyl transferase or alanine transaminase (i.e., if all three of these markers are elevated, then liver or bile duct injury is indicated), making it difficult to interpret this result.

We hypothesized that metBMI error could be leveraged to reveal perturbations in clinical chemistries that were not apparent in our primary analyses of antibiotic recency. Previously, we demonstrated that metBMI is more sensitive to lifestyle changes than traditional measures of BMI and that the error was associated with clinically relevant differences in metabolic health ^64^. Four of the 34 antibiotic-responsive metabolites are also metBMI features retained in at least one of the 10 LASSO models used to predict metBMI: 3-(4-hydroxyphenyl)lactate, alpha-hydroxyisocaproate, hydantoin-5-propionate, and indolepropionate. We found a significant interaction between metBMI error and recent antibiotic use on 25(OH)D levels (25-hydroxyvitamin D). Levels of 25(OH)D decreased as metBMI error increased, and this association was most pronounced in those with antibiotic use in the last three months (Fig. 5E), suggesting antibiotic-associated vitamin D depletion is only detectable in individuals with poor metabolic health. Vitamin D supplementation has been used to lower the risk of T2D in various cohorts, though its efficacy varied across populations ^90–92^. Moreover, conventionalization of germ-free mice was associated with increased levels of various vitamin D metabolites, including 25(OH)D ^93^. Interestingly, the same authors demonstrated an increase in 25(OH)D following antibiotic administration in vitamin D3-supplemented, germ-free mice ^94^, suggesting both indirect (i.e., microbiome-mediated) and direct mechanisms by which antibiotics may influence vitamin D homeostasis.

Lastly, we performed a large screen of metabolite-chemistry-antibiotic interaction effects, finding 20 instances where recent antibiotic use may moderate the association between metabolite and a clinical chemistry outcome. We found several instances where recent antibiotic use may be associated with a directional change in the relationship between metabolites and clinical chemistries. For instance, in recent antibiotic users, levels of tryptophan and 3-formylindole (a derivative of tryptophan) show positive associations with blood arsenic levels, though this association is negative in non-users. Antibiotic use has been associated with reduced fecal excretion of arsenic in mouse models ^95,96^, and in a small case-control study of Chinese adults chronically exposed to arsenic via drinking water, levels of tryptophan were negatively associated with arsenic-induced skin lesions ^97^. With the exception of one individual (whose blood arsenic was 31 mcg/L), levels of blood arsenic in the Arivale cohort were below the Quest Diagnostics reference range of less than 23 mcg/L ^98^. Thus, our findings are not directly comparable to studies on interventional or environmental arsenic exposure. However, these data reveal context-dependent associations between blood arsenic and tryptophan metabolites in individuals with recently perturbed gut microbiomes, in line with a recent study observing lower indolepropionate levels in arsenic-exposed mice with perturbed microbiomes ^99^. We also observed a moderation effect for the association between alpha-ketobutyrate and 3-methyl-2-oxobutyrate (also known as α-ketoisovalerate) with percentages of lymphocytes and neutrophils (Fig. 5G). BCAA metabolites have been implicated in T-cell activation and signaling ^100,101^, and circulating levels may play a role in connecting immune dysfunction with metabolic dysregulation ^102–104^. In a study investigating associations between whole-blood transcriptome modules and metabolic phenotypes, 3-methyl-2-oxobutyrate levels were positively associated with a neutrophil granule protein module ^103^. Interestingly, this module was negatively associated with diverse measures of health: fiber intake, insulin sensitivity, and indolepropionate levels ^103^. Taken together, these exploratory results highlight specific host physiological processes that appear to display context-dependent associations in the settings of recent antibiotic perturbations.

## Conclusion

In this study, we set out to resolve an apparent contradiction. On the one hand, many epidemiological studies that have focused on large-sized cohorts have indicated that antibiotic use increases T2D risk. On the other hand, several randomized, controlled intervention trials in small cohorts have not detected antibiotic-associated disruptions to host metabolic health ^18,19,39,54^. Most recently, Baldanzi et al. have analyzed the metagenomes of 14,979 individuals, identifying lasting changes in response to clindamycin, flucloxacillin and fluoroquinolone use ^2^. In many cases, antibiotic-enriched taxa were also associated with markers of cardiometabolic health, consistent with epidemiological studies linking antibiotic use with diabetes risk ^4,6,8^. We suspected that the contradiction between intervention studies and epidemiology stemmed from latent confounding and context-dependencies across cohorts. To investigate this hypothesis, we assembled multi-omic data from several cohorts, integrating gut microbiome, blood metabolome, blood proteome, clinical chemistry, diet, and lifestyle data to study the molecular fingerprint of antibiotic use across a wide variety of contexts. Overall, when controlling for a wide variety of covariates, we confirmed the interventional study results, showing no effect, on average, of antibiotic use on clinical chemistries. However, we found that individuals who were already metabolically vulnerable did see significant antibiotic-associated changes in clinical markers in the blood, suggesting vitamin defficiencies. Finally, we identified several gut bacterial genera that, when present, appeared to dampen or amplify the metabolomic impact of antibiotic use on the host. Future studies should investigate metabolic perturbation following antibiotic treatment with greater resolution of temporal, host dietary, and host environmental factors in order to inform prescribing standards and co-therapy design.

### Study Limitations

Our study has several limitations, notably the lack of complete antibiotic use metadata, including name, class, dosage, and indication. This may be partially mitigated by the recent observation that antibiotic-associated changes to gut microbiota composition tends to be very similar across different broad-spectrum antibiotic classes ^2^. In the current study, we cannot compare an individuals’ response and recovery to their pre-treatment state, given the lack of a pre-treatment sample. However, gut microbiome composition is largely stable over time ^105,106^, even following antibiotic treatment ^66^. Moreover, because individuals take antibiotics in response to a clinical indication, often an infection, our exposure variable necessarily includes that of the treated condition. Finally, we are always limited by unmeasured confounders in observational cohorts, but we were able to control for a long list of relevant covariates across our analyses. Despite these limitations, we successfully detect anticipated differences in gut microbiome diversity and composition following antibiotics (Fig. 1). We identified differences in several microbe-host co-metabolites following recent antibiotic treatment that were shown to be antibiotic-responsive in prior work ^34^, and we validated our results across several external cohorts.

## Methods

### Cohorts Arivale

The Arivale cohort consists of individuals aged 18 years or older, primarily from California and Washington, who self-enrolled in a former health wellness company (Arivale, Inc., USA). A detailed description of the cohort is provided in Zubair et al ^45^. Individuals who answered the recent antibiotic use question in the medication and health history questionnaires were considered for this study (n = 3,155). When asked about recent antibiotic use, answer choices were as follows: (0) “Within the last 3 months”; (1) “Within the last year”; or (2) “More than a year ago”. These choices were mutually exclusive. Thus, individuals who answered “within the last 3 months” do not also appear in the 12-month group. For this study, antibiotic use was coded as a categorical variable with three levels (“none”, “twelve”, and “three”; reference = “none”). Individuals missing covariate data (age, sex, BMI, medication usage, eGFR, and genetic ancestry) were excluded from this study (n = 489). Subsequent analyses include individuals from this subset of 2,666, provided their biosamples were collected within 30 days of their medication questionnaire. Procedures for this study were reviewed and approved by the Western Institutional Review Board, with institutional review board study numbers 20170658 for the Institute for Systems Biology and 1178906 for Arivale.

### Hispanic Community Health Study/Study of Latinos

HCHS/SOL is a prospective community-based cohort study of 16,415 self-identified Hispanic/Latino individuals living in 4 US metropolitan areas (Bronx, NY; Chicago, IL; Miami, FL; and San Diego, CA). A 2-stage area probability sample design was used to recruit participants (aged 18-74 at recruitment) and is described in more detail elsewhere. ^107,108^ The HCHS/SOL Gut Origins of Latino Diabetes (GOLD) ancillary study, conducted from 2016–2018, encompasses a subset of 3057 HCHS/SOL participants ^43^ and is approximately concurrent with the second in-person visit period (2014 to 2017), during which blood draws and clinical assessments were performed. In HCHS/SOL, individuals provided the date of last antibiotic use as month and year. The 15th of the month was imputed to calculate the time between antibiotic use and biosample collection. Individuals with a GOLD stool sample received within 60 days of Visit 2 (when blood is drawn for metabolomics) were included in the present study (n = 866). Antibiotic use was subsequently coded as a categorical variable with three levels (“none”, “six”, and “three”; reference = “none”). The study was approved by Institutional Review Boards at each field center where all participants gave written informed consent.

### MetaCardis Cohort

The MetaCardis cohort consists of adults from France, Denmark, and Germany enrolled in a prospective cross-sectional study investigating cardiac and metabolic phenotypes and has been described in detail elsewhere ^44^. Publicly available data from two studies were merged for analyses conducted in Figures 3A and 3B. Briefly, individual log-transformed serum metabolite data were obtained from Table S12 provided in Fromentin et al. ^44^. Individual-level metadata, anti-lipid and anti-diabetic medication use, antibiotic use in the last 5 years, and microbial load data were taken from Forslund et al. ^1^ Tables S1B, S2B, S2D, and Fromentin et al. Table S10, respectively.

### Gut Microbiome Sequencing

#### Arivale

Two external providers performed amplification and library preparation, with 300-bp paired-end MiSeq profiling of the 16S V3 + V4 region (DNAgenotek) or 250-bp paired-end MiSeq profiling of the 16S V4 region (Second Genome). Further analysis was performed using the denoise workflow from mbtools (https://github.com/gibbons-lab/mbtools) that wraps DADA2 ^109^. Taxonomy assignment was performed using the RDP classifier ^110^ with the SILVA database (version 132) ^111^. Here, 99% of the reads could be classified at the family level, 89% at the genus level, and 32% at the species level. A total of 1,967 individuals (out of 2,666) provided stool samples within 30 days of completing the antibiotic usage question.

#### HCHS/SOL

HCHS/SOL participants participating in the GOLD ancillary study were provided with a disposable paper inverted hat stool collection kit (Protocult collection device, ABC Medical Enterprises, Inc.). DNA extraction from fecal samples followed the Earth Microbiome Project protocol ^112^ and has been described in detail elsewhere ^43^. Shotgun sequencing was conducted in the Knight laboratory at the University of California San Diego (UCSD) using Illumina NovaSeq platforms ^113^. Prior to the CAZyme analysis, performed separately from the other microbiome analyses, raw shallow shotgun reads were trimmed of adapters and host contamination removed using kneaddata (including trimmomatic ^114^, bowtie2 human genome ^115^, and fastqc ^116^).

### Gut Microbiome Analysis

#### Arivale

For cross-sectional analyses, the “rrarefy” function in Vegan (v2.7.1) ^117^ was used to generate a count table rarefied to 21,869 reads. This count table was used for diversity calculations, logistic regression analysis (Fig. 2), and microbiome-metabolite moderation analysis (Fig. 4). Three diversity metrics were calculated as follows: (1) Richness: number of observed genera; (2) Dominance (Berger-Parker): number of reads assigned to the most abundant genus in a sample divided by the total number of reads in that sample; (3) Information (Shannon): *H* = 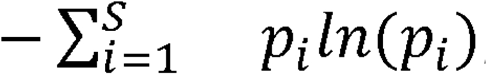, calculated using the “diversity” function in Vegan (v2.7.1). Principal coordinate analysis was performed using an ASV-level Bray-Curtis dissimilarity matrix rarefied to 21,869 reads (using the “avgdist” function in Vegan). Functional prediction (i.e., the number of carbohydrate-accessible enzyme families) from amplicon sequence variants was performed using Picrust2 ^50,51^ (v.2.6.0), which uses EPA-ng ^118^ and gappa ^119^ for placement of phylogenetic reads, castor ^120^ for hidden-state prediction, and MinPath ^121^ for pathway inference. Longitudinal assessment of gut microbiome richness was performed as above (albeit rarefied to 21,780 reads) for a subset of individuals who provided stool samples at least 60 days after the first sample (n = 837). Following rarefication, we removed participants with missing sequencing vendor (n=2), missing BMI (n=13), or with unexplained low reads (n=31 [all from one sequencing vendor]) at follow up, leaving 791 participants for the longitudinal analysis. To predict SCFA production, genus-level abundances were used to construct MCMMs using the community-scale metabolic modeling platform, MICOM ^48^ v0.37.1, as described previously ^49^. A presence/absence count table was created from our rarefied count table, where any non-zero count was converted to “1”. From here, taxa with prevalence between 3.5% and 97.5% were considered for testing. Taxa observed in only one sequencing vendor were removed (n=4), leaving n=209 taxa for analysis. Prior to the centered log-ratio (CLR) transformation of relative abundances, zero counts were imputed with half of the smallest non-zero observed count across the dataset. Subsequently, imputed values were reverted to missing (NaN) to avoid interpreting them as true observations. Finally, we applied prevalence filtering, retaining only the taxa present in at least 10% of samples.

#### HCHS/SOL

Of the 866 HCHS/SOL participants, 859 had available data in an OTU table matching the stool sample questionnaire. To exclude low-quality samples, those with <100000 reads were removed (n = 5), leaving 854 participants with antibiotic recency data, stool shallow shotgun metagenomic data, and for which stool samples were returned within 60 days of HCHS/SOL V2 (blood for metabolomics taken at V2). Individuals with missing BMI (n = 3) and eGFR (n = 5) data were removed, leaving n = 846 for microbiome diversity analyses. 2709 species were detected in these samples. These were agglomerated to the genus level for harmonization with Arivale using the “tax_glom” function in the phyloseq R package ^122^ (v 1.50.0), to 868 genera. Alpha diversity calculations and microbiome analyses were subsequently performed using the same approaches and prevalence cut-offs listed above. For the CAZyme analysis in HCHS/SOL using Cayman ^52^, the human-gut database was downloaded from Zenodo (https://zenodo.org/records/10473258?preview_file=gene_catalogues.zip) and indexed with BWA, and the human-gut CAZyme annotation file also downloaded for use with the Cayman tool. Clean paired reads along with any orphaned reads were then processed using the Cayman profile function. For this analysis, raw reads were available from 822 participants (646 with no antibiotic use in the past 6 months, 73 with use 3-6 months prior to stool sample, and 135 within 0-3 months). CAZyme richness was calculated by counting the number of uniquely observed CAZyme families within each sample, provided that the abundance was > 1 RPKM ^52^.

### Enterotype Analysis

Dirichlet Multinomial Mixture (DMM) modeling ^53^ was performed on rarefied genus-level count tables for both Arivale and HCHS/SOL cohorts. To select the optimal number of enterotype clusters, we used the Bayesian information criterion (BIC) which resulted in 4 enterotypes for Arivale and 3 for SOL. Logistic regression models were used to test enrichment of *Bacteroides* enterotypes among antibiotic users. In Arivale, this was specified by testing for “Bac2” versus any other enterotype, given the health relevance of this specific enterotype. In HCHS/SOL, the 3 and 6 month groups were combined to a single antibiotic use group, and “Bac” versus any other enterotype was tested.

### Plasma and Serum Metabolomics

Metabolomic data for the cohorts included in Figure 3A were generated by Metabolon Inc. (Durham, NC) using methods described in detail elsewhere ^44,123^. Untargeted metabolomics was performed on either plasma (Arivale) or serum (HCHS/SOL, MetaCardis) samples. Briefly, all measurements were taken using Waters ACQUITY ultraperformance liquid chromatography (UPLC) and a Thermo Scientific Q-Exactive high-resolution/accuracy mass spectrometer interfaced with a heated electrospray ionization (HESI-II) source and an Orbitrap mass analyzer operated at 35,000 mass resolution. Four sample fractions (including one backup) underwent measurement using different approaches. The first aliquot was analyzed under positive-ion conditions by elution from a C18 column using water and methanol with 0.05% perfluoropentanoic acid and 0.1% formic acid. The second aliquot additionally used acetonitrile. The third aliquot was analyzed under negative-ion optimized conditions and gradient eluted from a C18 column using methanol, water, and 6.5 mM ammonium bicarbonate at pH 8.0. The final aliquot was measured using negative ionization following elution from a HILIC column using water, acetonitrile, and 10 mM ammonium formate at pH 10.8. MS analyses utilized MS and data-dependent multistage MS scans with dynamic exclusion at an approximate range of 70–10,000 m/z. Metabolites not meeting annotation criteria but consistently detected in various biofluids are labeled with the prefix ‘X’ followed by a unique ID. For Arivale, the discovery cohort, metabolites observed in fewer than 30% of samples were excluded, leaving 969 metabolites for analysis. Metabolites were log-transformed and Z-scored prior to analysis. Finally, regression models were fit for each metabolite individually (dependent variable), including antibiotic recency as the predictor of interest and adjusting for all covariates included in the “full model”.

Results were subsequently validated in two external cohorts: HCHS/SOL and MetaCardis. Here, individual linear regression models were constructed for the 34 antibiotic-responsive metabolites (BH q < 0.1). For HCHS/SOL, each metabolite was regressed on antibiotic recency, with antibiotic recency as the predictor of interest, adjusting for age, sex, BMI, field center, and eGFR. For HCHS/SOL, metabolite data was available for 815 individuals, and three were removed due to missing BMI, leaving 812 participants for analysis. For MC, each metabolite was regressed on the predictor of interest—the total number of antibiotic courses in the last 5 years or binarized antibiotic use—adjusting for age, gender, BMI, microbial load, study center, antidiabetic medication use (encoded as 0 or 1), and antilipid medication use (encoded as 0 or 1). To compare the effect of antibiotics versus glucose intolerance on the 34 antibiotic-responsive metabolites, data from MC were re-analyzed as raw data were publicly available, and significant findings from 3 cohorts (Qatari ^59^, Swedish ^60^, HCHS/SOL ^61^) were compiled and merged to produce Figure 3B. To re-analyze MC data, each metabolite was regressed on health status as the predictor of interest, adjusting for age, gender, BMI, microbial load, study center, antidiabetic medication use, antilipid medication use, and the total number of antibiotic courses in the last 5 years. Significant findings were obtained from Yousri et al. Tables S2, Wu et al. Table S8, and Li et al. Table S2. Compiled data used to create Fig. 3B is provided in Table S6.

### Arivale Plasma Proteomics

Plasma protein levels for Arivale were measured using Proseek Inflammation, Cardiovascular II, and Cardiovascular III panels (Olink Biosciences). Data was processed and batch corrected as described previously ^124^. Data was matched to antibiotic recency questionnaire data with a threshold of 30 days. We applied a 70 percent prevalence filtering threshold, resulting in 274 proteins for analysis.

### Arivale Metabolic BMI Prediction Analysis

Beta coefficients from ten sparse metabolomics-inferred BMI (metBMI) models previously trained on Arivale metabolomic data using LASSO regression ^64^ were used to generate metBMI predictions for the antibiotic recency cohort (as individuals having used antibiotics in the last 3 months were excluded in Watanabe et al.). Outputs from the ten models were averaged to produce the final metBMI prediction. Correlation between measured and metabolic BMI was assessed using Pearson’s correlation. Error was calculated as metBMI minus measured BMI (kilograms/meters^2^), producing an error term used in subsequent analysis. Finally, individual linear regressions were performed for 94 clinical chemistries, using the following model: chemistry ∼ error * abx_recency + BMI:abx_recency + BMI:error + covariates. A BMI:error interaction term was included because metBMI error prediction is known to vary by BMI (for example, underprediction is more common in high BMI ranges).^64^ However, the predictor of interest in these analyses is a two-way interaction between error and antibiotic recency.

### Arivale Dietary Questionnaire Data

Response rates for self-reported questionnaires in Arivale vary. Analysis of dietary variables was limited to dietary variables with at least 100 responses, yielding a set of 12 dietary variables: diet type (no special diet, vegetarian, diabetic, etc.), alcohol preference (no preference, beer, wine, or liquor), alcohol days per week, alcohol drinks per day, breakfast frequency, snack frequency, cruciferous vegetable frequency, fruits per day, vegetables per day, sugary drinks per day, grains per day, and cups of water per day. Two were removed due to small sample sizes (<10 in at least one cell): diet type and cups of water per day. Subsequently, all dietary variables except alcohol preference were binarized to reflect regular or near-daily consumption. Finally, we retained alcohol drinks per day rather than alcohol days per week, as the two capture largely overlapping information following binarization, leaving 9 dietary variables for analysis.

### Statistical analysis

Data management was performed using either pandas ^125,126^ (v2.3.3) in Python (Jupyter Notebooks) or R (RStudio). Unless otherwise specified, the full model used in Arivale linear regressions adjusts for the following covariates: age, sex, BMI, genetic ancestry principal components 1-5, blood sugar medication use (“Yes” versus “No”), cholesterol-lowering medication use (“Yes” versus “No”), and eGFR. Values for time-varying covariates were obtained from questionnaires collected at the beginning of individuals’ participation in the Arivale program. eGFR was calculated based on the CKD Epidemiology Collaboration (CKD-EPI) creatinine Equation, as recommended by the current guidelines of the National Kidney Foundation ^127^: eGFRcr = 142 x min(Scr/k,1) x max(Scr/k,1)^−1.200^ x 0.9938^Age^ x 1.012 [if female], where Scr = standardized serum creatinine in mg/dL, k = 0.7 (female) or 0.9 (male), and = −0.241 (female) or −0.302 (male). History of T2D was defined as a self-report of current or past T2D. Statistical analyses for cross-sectional regressions were performed using statsmodels ^128^ (v0.14.6). For linear regressions, hypotheses were tested for significance with two-sided t-tests. The presence or absence of microbial taxa was modeled using logistic regression with hypotheses tested using Wald Z-tests. All tests were FDR-adjusted using the Benjamini–Hochberg ^129^ method (q < 0.1). The “check_collinearity” function in the R performance package ^130^ (v0.16.0) was used to assess multicollinearity in both simple and mixed linear models. Analysis of longitudinal changes in gut microbiome alpha diversity between antibiotic recency groups was assessed using linear mixed models with a random intercept in the lme4 R package ^131^ (v1.1.37). Individual models were created for observed features, Berger-Parker, and Shannon index. In these analyses, all covariates except for time, sequencing vendor, and BMI were obtained from the baseline intake questionnaires and were treated as time-invariant. Time was calculated as the difference in days between stool samples and subsequently divided by 100. Follow-up BMI measurements were used to calculate within-person (*BMI_w_* = *BMI_t,i_* 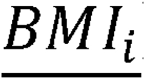) and between-person (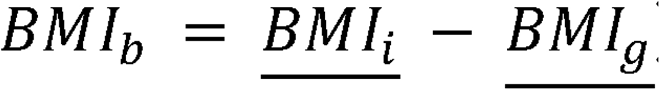) BMI variables, where 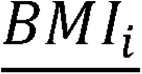, denotes a BMI measurement for individual i at time t, denotes the mean of two BMI measures for individual i, and 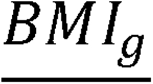 denotes the grand mean across all participants and time points. Baseline alpha diversity measures were not included in the model. Because stool samples were collected near the medication questionnaire date, baseline diversity is a consequence of the reported antibiotic use rather than a pre-exposure characteristic. To test for an interaction between change in alpha diversity over time and antibiotic recency, a likelihood ratio test (ML estimation) was used to compare the covariate-adjusted full model with a reduced model lacking the interaction between time and antibiotic recency. Group-specific slopes and pairwise slope contrasts were estimated post hoc using emmeans ^132^ (v2.0.3) (Kenward–Roger df, Tukey adjustment).

## Data availability

Qualified researchers can access the full Arivale deidentified dataset supporting the findings in this study for research purposes through signing of a data use agreement. Inquiries to access the data can be made at and will be responded to within seven business days. The raw 16S amplicon sequencing data for Arivale are available in the Sequencing Read Archive under BioProjects: PRJNA826530 and PRJNA826648.

The data and computer code used for the HCHS/SOL analyses can be obtained through the cohort study’s coordinating center at University of North Carolina-Chapel Hill. HCHS/SOL permits data sharing for HCHS/SOL–approved manuscript proposals with outside investigators. All data sharing is conducted in accordance with HCHS/SOL study and NIH policies and governed by a Data and Materials Distribution Agreement (DMDA) between UNC and the external institution, ensuring the confidentiality and privacy of HCHS/SOL participants and their families. Alternatively, de-identified HCHS/SOL data are publicly available at BioLINCC and dbGaP for the subset of the study cohort who authorized general use of their data at the time of informed consent.

## Code availability

The code used in this study is available at: https://github.com/Gibbons-Lab/2026_cperez_antibiotic_recency

## Contributions

Conceptualization was carried out by C.P. and S.M.G. Methodology was the responsibility of C.P. and S.M.G. and C.P., S.K.A., L.P., and N.Q-B. performed formal analyses. Data and code curation was performed by C.P. Resources and tools were provided by L.P., N.Q-B., A.T.M., N.R., and S.M.G. Visualizations were produced by C.P. The original draft was written by C.P. and all other authors were involved in editing and approving the final manuscript. Supervision and project administration was provided by N.R., R.K., and S.M.G.

## Data Availability

Qualified researchers can access the full Arivale deidentified dataset supporting the findings in this study for research purposes through signing of a data use agreement. Inquiries to access the data can be made at and will be responded to within seven business days. The raw 16S amplicon sequencing data for Arivale are available in the Sequencing Read Archive under BioProjects: PRJNA826530 and PRJNA826648.
The data and computer code used for the HCHS/SOL analyses can be obtained through the cohort study's coordinating center at University of North Carolina-Chapel Hill. HCHS/SOL permits data sharing for HCHS/SOL-approved manuscript proposals with outside investigators. All data sharing is conducted in accordance with HCHS/SOL study and NIH policies and governed by a Data and Materials Distribution Agreement (DMDA) between UNC and the external institution, ensuring the confidentiality and privacy of HCHS/SOL participants and their families. Alternatively, de-identified HCHS/SOL data are publicly available at BioLINCC and dbGaP for the subset of the study cohort who authorized general use of their data at the time of informed consent.
All other data sets used in this study were publicly available.
The code used in this study is available at:
https://github.com/Gibbons-Lab/2026_cperez_antibiotic_recency

## Acknowledgments

Research reported in this publication was supported by the National Institute of Diabetes and Digestive and Kidney Diseases (R01DK133468 [to S.M.G.], R01DK119268, R01DK143633, and 5R01DK134672) and the National Institute on Aging (U19AG023122 and R01AG085320) of the National Institutes of Health (NIH). S.K.A. received support from National Cancer Institute T32CA094880 during this work. The Hispanic Community Health Study/Study of Latinos is a collaborative study supported by contracts from the National Heart, Lung, and Blood Institute to the University of North Carolina (75N92025D00001, HHSN268201300001I/N01-HC-65233), University of Miami (75N92025D00009, HHSN268201300004I/N01-HC-65234), Albert Einstein College of Medicine (75N92025D00007, HHSN268201300002I/N01-HC-65235), University of Illinois at Chicago (75N92025D00008, HHSN268201300003I/N01-HC-65236 Northwestern Univ), and San Diego State University (75N92025D00010, HHSN268201300005I/N01-HC-65237). The authors thank the staff and participants of HCHS/SOL for their important contributions. The content is solely the responsibility of the authors and does not necessarily represent the official views of the NIH. The funders had no role in designing, carrying out or interpreting the work presented in the manuscript. The following figures were created in https://BioRender.com: Fig. 1A, 1E, 4E, 4D, 5B, 5F, and S1.

