## Supplemental Figures 1-4 for "Context-dependent associations between recent antibiotic treatment, gut microbiome composition, blood metabolites, and metabolic health"

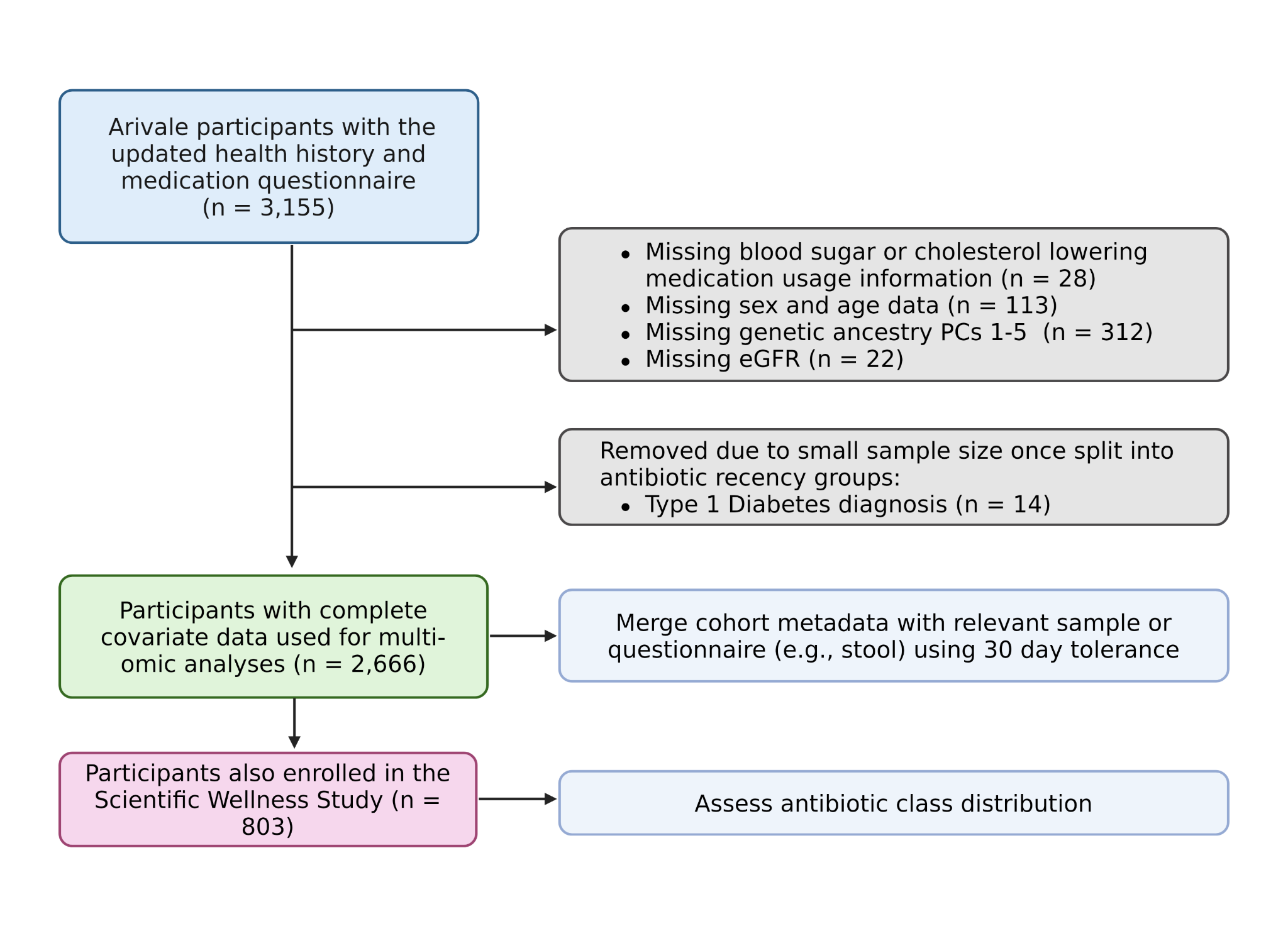

**Supplementary Figure 1. Arivale cohort flow diagram.**

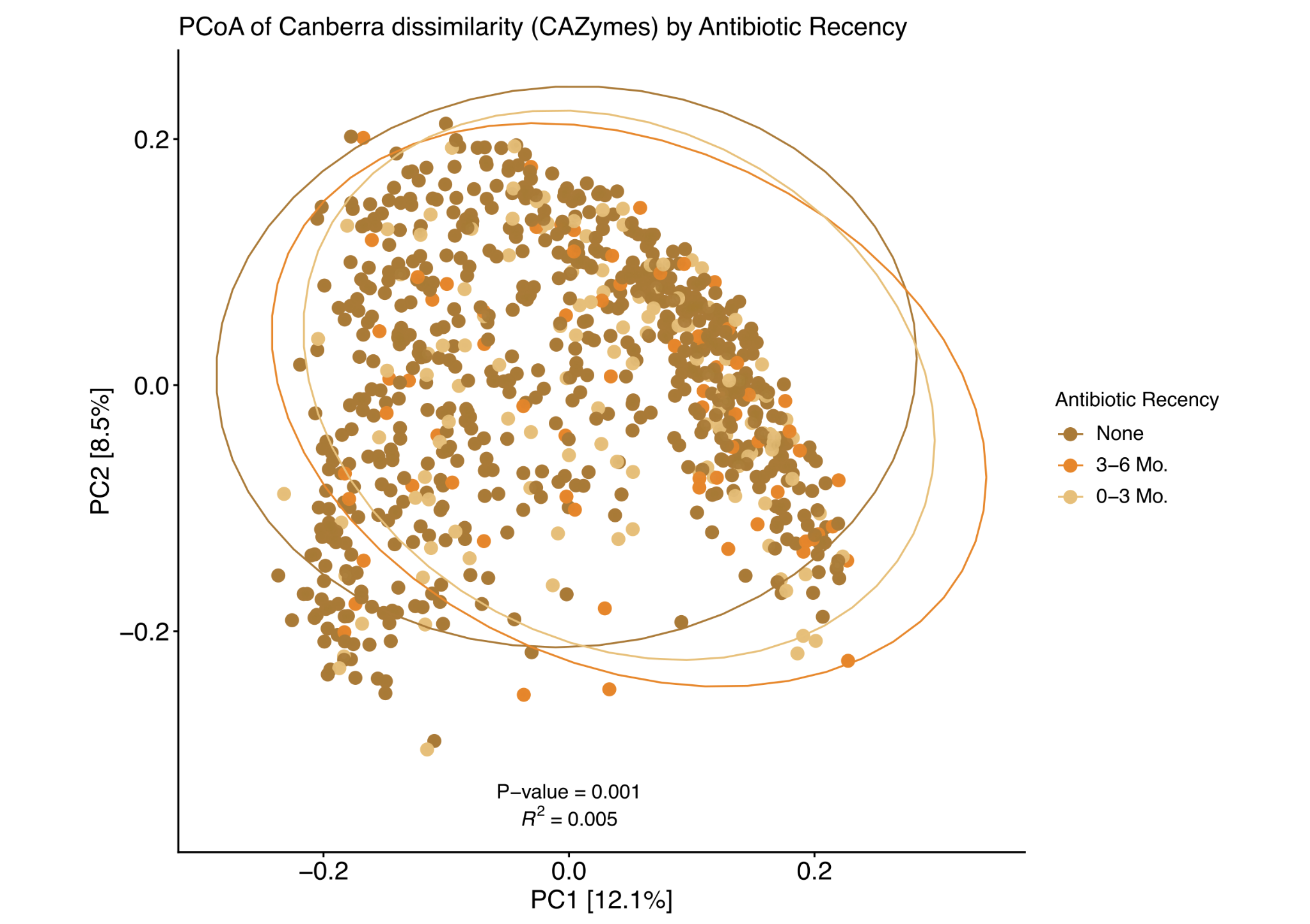

**Supplementary Figure 2. Recent antibiotic use is associated with global changes in the CAZyme repertoire.** Principal coordinate analysis using Canberra dissimilarity of CAZyme families by antibiotic recency. P-value and R^2^ from the unadjusted model are shown on the figure (PERMANOVA, R^2^ = 0.005, p = 0.001). PERMANOVA adjusted for age, sex, field center, BMI, eGFR, and birthplace (US Born or not) showed similar results (R^2^ =0.004, p = 0.002).

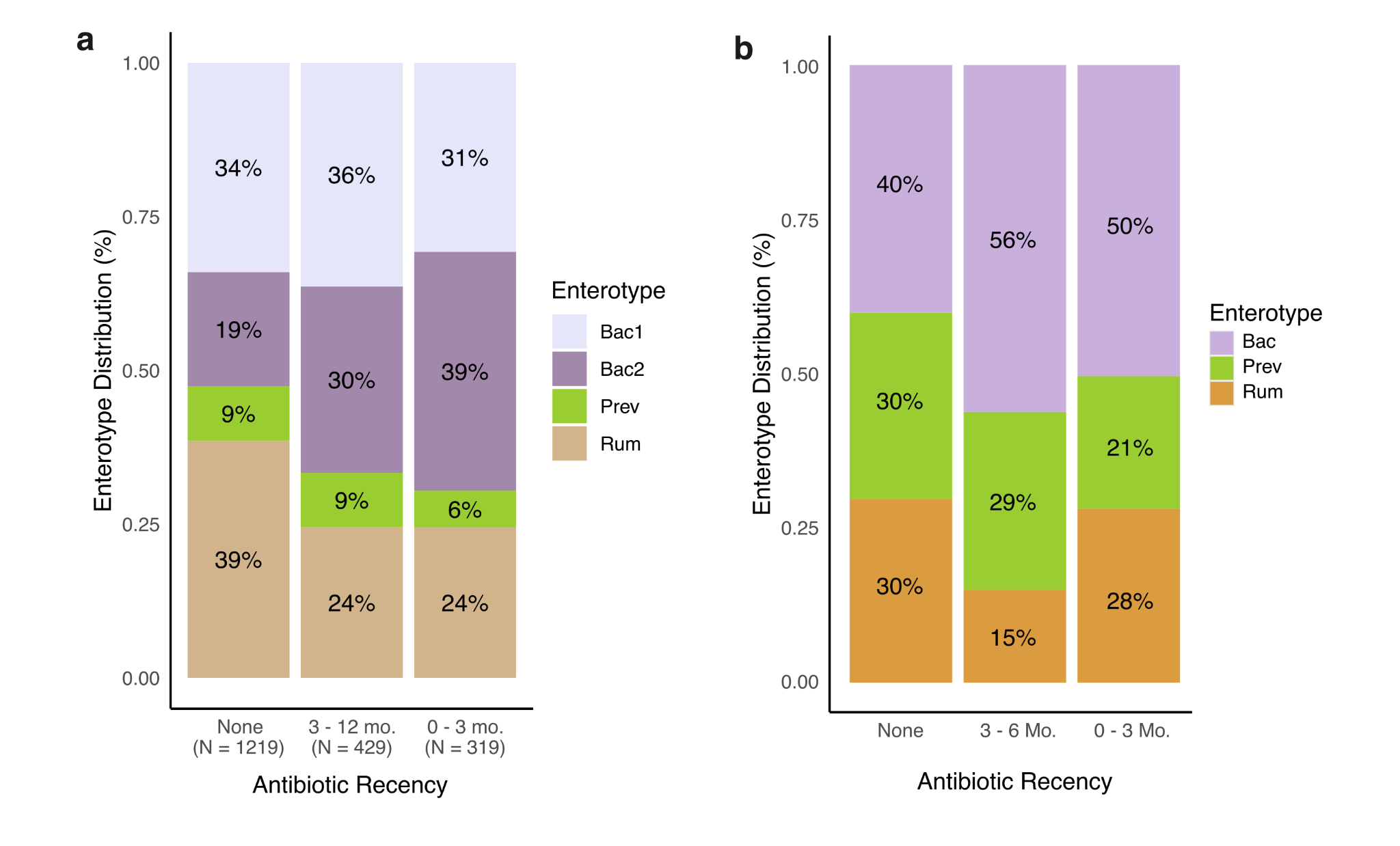

**Supplementary Figure 3. Recent antibiotic use is associated with enrichment for Bacteroides enterotypes.** Stacked bar chart displaying the distribution of enterotypes by antibiotic recency groups within **a**, Arivale and **b**, SOL. Bac2 enterotypes are enriched among recent antibiotic users in Arivale (OR_3_ = 2.89, BH q = 6.96 x 10^-13^; OR_12_ = 1.75, BH q = 4.04 x 10^-05^), while the Rum enterotype is depleted (OR_3_ = 0.48, BH q = 1.23 x 10^-06^; OR_12_ = 0.54, BH q = 9.62 x 10^-06^). A similar enrichment of Bac is observed in the SOL cohort (OR = 1.53 , p = 0.01). Coefficients were tested using two-sided Wald tests. Arivale tests were corrected for multiple hypotheses testing (Bac2 and Rum outcomes) using BH (FDR q < 0.1).

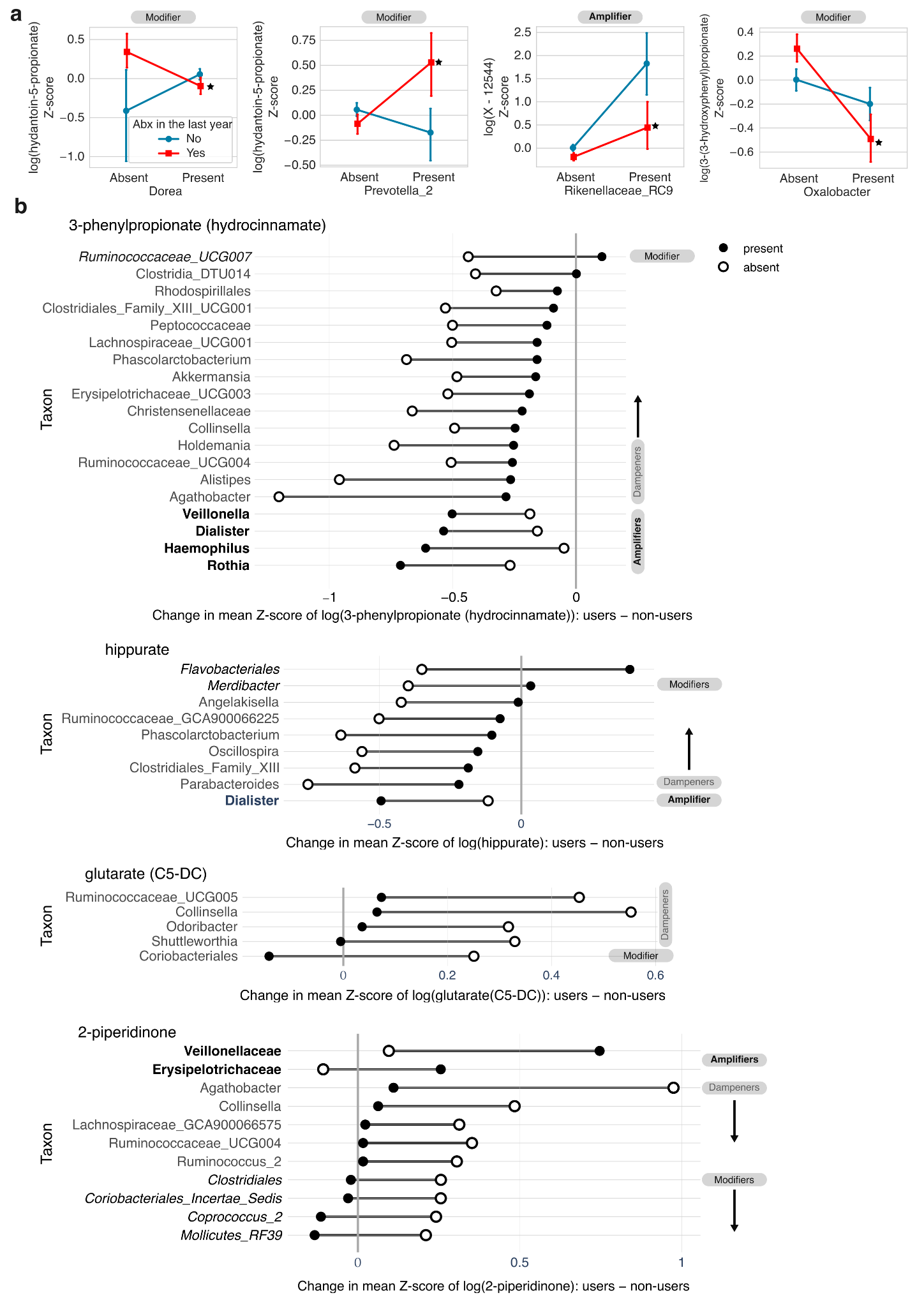

**Supplementary Figure 4. Moderation of blood metabolites by presence of microbial genera.** **a**, Point plots displaying the change in normalized blood metabolite abundance in antibiotic users (red) and non-users (blue). Metabolites from left to right: hydantoin-5-propionate (left and middle left), X-12544 (middle right), and 3-(3-hydroxyphenyl)propionate (right). Bacterial genera from left to right: *Dorea*, *Prevotella_2*, *Rikenellaceae_RC9*, and *Oxalobacter*. **b**, Barbell plot showing the difference in mean normalized blood metabolite levels between antibiotic users and non-users for metabolites with 5 or more potential moderators (interaction term BH q <0.1 for all genera shown). Metabolites from top to bottom: 3-phenylpropionate (hydrocinnamate), hippurate, glutarate (C5-DC), and 2-piperidinone. Genera considered “amplifiers” are denoted in bold, “modifiers” are italicized in black, and “dampeners” are in grey normal typeface. Closed and open circles represent genus presence or absence, respectively. For point plots, stars indicate significant interaction effect with FDR q-value thresholds < 0.1.
